# Associations of Device-Measured Sleep Duration, Regularity, and Efficiency with Cardiometabolic Health in Adults: Findings from the ProPASS Consortium

**DOI:** 10.1101/2025.07.22.25332021

**Authors:** Annemarie Koster, Raaj Kishore Biswas, Matthew N Ahmadi, Joanna M Blodgett, Nicholas A Koemel, Wenxin Bian, Andrew J Atkin, Richard M Pulsford, Borja del Pozo Cruz, Carlos Celis-Morales, John Mitchell, Pasan Hettiarachchi, Peter J Johansson, Magnus Svartengren, Hsiu-Wen Chan, Kristin Suorsa, Esmée A Bakker, Sari Stenholm, Thijs M.H. Eijsvogels, Hans HCM Savelberg, Vegar Rangul, Alun D Hughes, I-Min Lee, Peter A. Cistulli, Jean-Philippe Chaput, Andreas Holtermann, Mark Hamer, Emmanuel Stamatakis, ProPASS Collaboration

**Affiliations:** Department of Social Medicine, CAPHRI Care and Public Health Research Institute, Maastricht University, Maastricht, the Netherlands; Mackenzie Wearables Research Hub, Charles Perkins Centre, University of Sydney, Sydney, NSW, Australia; School of Health Sciences, Faculty of Medicine and Health, University of Sydney, Sydney, NSW, Australia; Institute of Sport Exercise and Health, Division of Surgery and Interventional Sciences, UCL, United Kingdom; University College London Hospitals NIHR Biomedical Research Centre, London, UK; School of Health Sciences, University of East Anglia, Norwich, UK; Faculty of Health and Life Sciences, University of Exeter, Exeter, UK; Department of Sport Sciences, Faculty of Medicine, Health, and Sports, Universidad Europea de Madrid, Madrid, Spain; School of Cardiovascular and Metabolic Health, University of Glasgow, Glasgow, United Kingdom; Occupational and Environmental Medicine, Department of Medical Sciences, Uppsala University, Uppsala, Sweden; Australian Women and Girls’ Health Research Centre, School of Public Health, The University of Queensland, Brisbane, Queensland, Australia; Department of Public Health, University of Turku and Turku University Hospital, Turku, Finland; Centre for Population Health Research, University of Turku and Turku University Hospital; Turku, Finland; Department of Primary and community care, Radboud university medical center, Nijmegen, the Netherlands; Department of Medical BioSciences, Exercise Physiology Research Group, Radboud University Medical Center, Nijmegen, the Netherlands; Department of Nutrition and Movement Science, NUTRIM School for Nutrition and Translational Research in Metabolism, Maastricht University, Maastricht, the Netherlands; HUNT Research Centre, Department of Public Health and Nursing, Faculty of Medicine and Health Sciences, Norwegian University of Science and Technology (NTNU), Levanger, Norway; MRC Unit for Lifelong Health and Ageing, UCL Institute of Cardiovascular Science, UCL, United Kingdom; UCL BHF Research Accelerator, University College London, London, UK; Division of Preventive Medicine, Brigham and Women’s Hospital and Harvard Medical School, Boston, Massachusetts, USA; Department of Epidemiology, Harvard TH Chan School of Public Health, Boston, Massachusetts, USA; Sleep Research Group, Charles Perkins Centre, University of Sydney, Camperdown, New South Wales, Australia; Northern Clinical School, Faculty of Medicine and Health, University of Sydney, Camperdown, New South Wales, Australia; Department of Respiratory and Sleep Medicine, Royal North Shore Hospital, St Leonards, New South Wales, Australia; Healthy Active Living and Obesity Research Group, Children’s Hospital of Eastern Ontario Research Institute, Ottawa, Ontario, Canada; Department of Pediatrics, University of Ottawa, Ottawa, Ontario, Canada; National Research Centre for the Working Environment, Copenhagen, Denmark

**Author notes:** Joint first authors (Annemarie Koster, Raaj Kishore Biswas). Joint senior authors (Mark Hamer, Emmanuel Stamatakis). ProPASS collaboration: Carla JH van der Kallen: Department of Internal Medicine, CARIM School for Cardiovascular Diseases Maastricht University, Maastricht, the Netherlands. Dick HJ Thijssen: Department of Medical BioSciences, Exercise Physiology research group, Radboud university medical center, Nijmegen, the Netherlands Nidhi Gupta: National Research Centre for the Working Environment, Copenhagen, Denmark Lauren B Sherar: School of Sport, Exercise and Health Sciences, Loughborough University, UK. Corresponding author: Annemarie Koster, PhD.

**Keywords:** cardiometabolic health, individual participant meta-analysis, sleep duration, sleep efficiency, sleep regularity, wearables

## Abstract

**Background:** Sleep is critical for cardiometabolic health, yet evidence on the independent and combined associations of device-measured sleep parameters remains limited. The aim of this study is to examine the independent and joint associations of sleep duration, regularity, and efficiency with cardiometabolic health outcomes in adults.

**Methods:** Cross-sectional data from six cohorts (n=14,085 participants; five countries, ≥4-day wear time) from the Prospective Physical Activity, Sitting and Sleep (ProPASS) consortium were used. Sleep duration (short: <7 h/day; adequate: 7-8 h/day; long: >8 h/day), sleep regularity index as a measure of day-to-day variability in sleep-wake patterns (regular: >87.3%; slightly irregular: 71.6-87.3%; irregular: <71.6%), and sleep efficiency as the ratio of total sleep time to total time in bed (high: >91.8%), medium: 85.3-91.8%, low: <85.3%; based on tertiles) were derived from thigh-worn accelerometer data. Cardiometabolic health markers included body mass index, waist circumference, HDL and LDL cholesterol, glycated haemoglobin, systolic and diastolic blood pressure, and a composite cardiometabolic risk z-score was computed. Generalized linear regression was used to examine individual and joint associations of sleep parameters with cardiometabolic health, adjusted for age, sex, cohort, smoking, alcohol consumption, medication use, prevalent cardiovascular disease and moderate-to-vigorous intensity physical activity.

**Results:** Short sleep duration (β: 0.05, 95%CI: 0.03-0.07), an irregular sleep pattern (β:0.14, 95%CI:0.11-0.18), and low sleep efficiency (β:0.10, 95%CI:0.07-0.13) were associated with a higher cardiometabolic risk z-score compared to adequate sleep duration, regular sleep patterns, and high sleep efficiency, respectively. A long sleep duration was not associated with cardiometabolic risk score (β:0.01, 95%CI:-0.03-0.03). The joint analysis of all three sleep parameters shows that individuals with an irregular and low sleep efficiency, regardless of sleep duration, had worse cardiometabolic health (in long sleepers: β:0.21, 95%CI: 0.15-0.28; in adequate sleepers: β:0.23, 95%CI: 0.16-0.30; in short sleepers: β:0.28, 95%CI: 0.22-0.35) compared to adequate, regular and highly efficient sleepers.

**Conclusion:** Our study suggests that sleep regularity and efficiency are important for cardiometabolic health beyond sleep duration. Future longitudinal studies and trials should evaluate multidimensional sleep indices that incorporate duration, regularity, and efficiency across diverse populations.

## Introduction

Sleep is widely recognized as critical for maintaining overall health. ^1^ There is established evidence for associations between sleep duration and health outcomes including cardiometabolic conditions such as type 2 diabetes, excess adiposity and high blood pressure.^1^ Typically, studies report a U-shaped association between sleep duration and health, indicating that both shorter and longer sleep duration are associated with increased risk of mortality and cardiometabolic disease risk, including obesity, diabetes and cardiovascular disease.^2–4^ While most research has focused on sleep duration, more qualitative aspects of sleep have also shown to be independently associated with health. For example, a meta-analysis of 17 prospective cohort studies showed that lower self-reported sleep quality (defined as difficulty maintaining sleep or disturbed sleep) has been associated with coronary heart disease risk but not with other cardiovascular outcomes or mortality.^5^ Poor sleep efficiency (i.e. spending a large proportion of time in bed awake rather than asleep), quantified by in-home polysomnography, has also been associated with increased cardiovascular disease risk.^6^

In addition to sleep duration and efficiency, emerging evidence suggests that the stability of sleep-wake cycles is also important for health. ^7^ The variability of sleep patterns, particularly the duration and timing of sleep, has the potential to disrupt the circadian rhythm of biological processes. Such disruption is known to, for example, increase cardiovascular risk.^8^ A systematic review examining the associations of sleep timing and sleep regularity with health outcomes in adults reported that higher sleep variability was associated with adverse health outcomes.^7^ However, the evidence was rated as very low to moderate quality, primarily due to reliance on self-reported sleep behaviours, which are subject to reporting bias and limit the ability to translate findings into public health guidance.^7^ A systematic review of sleep variability and cardiometabolic health shows that variability is likely associated with obesity, weight gain, cardiovascular disease, and metabolic syndrome but not with insulin resistance.^9^ It is important to note that many studies that have examined associations between sleep variability and health have not adjusted for sleep duration. More recently, findings from the Multi-Ethnic Study of Atherosclerosis have shown that a greater device-measured sleep irregularity was cross-sectionally associated with obesity, hypertension, fasting glucose, and haemoglobin A1C^10^, and prospectively associated with metabolic syndrome^11^ and incident cardiovascular disease.^12^ A prospective study from the Hispanic Community Health Study/Study of Latinos found no association between device-measured sleep regularity and glucose biomarkers or incident diabetes^13^. Recent studies, using data from the UK Biobank begun, exploring the joint associations of sleep duration and sleep regularity, showing that high sleep irregularity was associated with an increased risk of type 2 diabetes risk and major adverse cardiovascular events among people who achieved the recommended sleep duration.^8,14^ This evidence suggests that other sleep domain beyond sleep duration alone, may affect health.

Most epidemiological studies to date have relied on self-reported sleep characteristics, which have several limitations; in contrast, device-based assessments offer greater validity and higher data resolution.^15^ Further, the use of 24-hour assessment protocols is especially appealing for the assessment of sleep regularity over multiple nights.^16^ Wrist-worn accelerometers are commonly used for sleep assessment, and validation studies comparing them to polysomnography, the gold standard for sleep measurement, have shown that while they are highly sensitive in detecting sleep, their specificity in identifying wakefulness is relatively low.^17^ Recently, Johansson et al. ^18^ have shown that sleep duration can be measured with comparable accuracy using a thigh-worn accelerometer. Using data from the Prospective Physical Activity, Sitting and Sleep (ProPASS) consortium, we examined the independent and joint associations of sleep duration, regularity and efficiency with cardiometabolic health markers. This ProPASS resource consists of harmonized individual participant data of six cohorts from the Netherlands, UK, Australia, Denmark, and Finland with thigh-worn accelerometer data.

## Methods

### Consortium sample

ProPASS is an international research collaboration platform consisting of observational cohort studies of adults with thigh-worn accelerometry data.^19^ For this project, we included cross-sectional data from six participating studies that were pooled and harmonized: The Maastricht Study (TMS; The Netherlands, n=7273)^20^, the 1970 British Birth Cohort Study (BCS70; UK, n=4786)^21^, the Australian Longitudinal Study on Women’s Health (ALSWH; Australia, n=918)^22^,the Danish PHysical ACTivity cohort with Objective measurements cohort (DPhacto; Denmark, n=426)^23^, the Nijmegen Exercise Study (NES; The Netherlands, n=535)^24^, and the Finnish Retirement and Aging Study (FIREA; Finland, n=147)^26^. **Supplemental table 1** presents detailed information for each cohort. Data were physically pooled at the University of Sydney adhering to cohort-specific requirements and necessary data transfer agreements; this included harmonization of covariates and outcomes, as well as cleaning and processing of raw accelerometer data.^27,28^ Ethical approval and informed consent were obtained at the cohort level.

### Sleep

All cohorts collected movement behaviour data using a 7-day, 24 h/day thigh-worn accelerometer protocol; four studies used ActivPAL devices (BCS70, TMS, ALSWH, NES), one used Axivity devices (FIREA), and one used ActiGraph devices (DPhacto). Raw accelerometer data were centrally processed using previously validated software, ActiPASS v 1.32^29^. ActiPASS identifies behaviours in 2 s windows with a 50% overlap, resulting in a resolution of 1 s epochs, and implements algorithms for non-wear, sleep detection, posture, and activity intensity.^18,30,31^ Validation against polysomnography has shown good sensitivity (0.84) and accuracy (0.80) to detect sleep.^18,31^ Participants with a minimum of four valid wear days (≥20 hours per day), including at least one weekend day were included in the analyses. We estimated sleep duration using a validated algorithm that identified thresholds for wakefulness and sleep.^18^ Sleep duration (h/day) was categorized into short (<7 h/day), adequate (7-8 h/day), and long (>8 h/day), based on evidence from an umbrella review that 8-8 hours per day is most favourably associated with health outcomes.^1^ Sleep efficiency (%) defined as the ratio of sleep period time to total time in bed, was subsequently categorized into tertiles (high: efficiency >91.8%; medium, 85.3%≤ efficiency ≤ 91.8%; low: efficiency<85.3%).^32^The Sleep Regularity Index (SRI, %) represents the percentage probability of an individual maintaining the same sleep/wake state on adjacent days, with a scale from 0 (totally irregular) to 100 (perfectly regular).^33^ Based on previous research using device data from the UK Biobank, we classified participants into regular (>87.3), slightly irregular (71.6-87.3), and irregular (<71.6) sleep regularity groups^8, 34^. Details on the sleep variables and source code are shown in **Supplemental Table 2**.

### Cardiometabolic health markers

Harmonized cardiometabolic health markers included body mass index, waist circumference, cardiometabolic blood markers, and blood pressure.^27^ Body mass index (BMI, kg/m^2^; calculated from height and weight) and waist circumference (cm) were assessed by trained nurses or researchers during home or clinic-based visits. Cardiometabolic blood biomarkers were measured in five studies (not available in DPhacto) and included: total and HDL cholesterol (mmol/L), triglycerides (mmol/L), and glycated haemoglobin (HbA1c, mmol/mol; measured in ALSWH, BCS70, and TMS only). LDL cholesterol was calculated based on the Friedewald equation.^35^ Measurement and assay methodology were similar across cohorts, with consistently low coefficients of variation (< 6%). Blood pressure measurement protocols were consistent across the six cohorts, each of which used automated blood pressure monitors administered by a research nurse or study staff member to assess systolic and diastolic blood pressure (mmHg). Details on the assessment of blood biomarkers and the measurements across cohort are presented in **Supplemental Table 3 and 4**. A composite cardiometabolic risk score was computed by averaging the standardized values (z-scores) of eight biomarkers: BMI, waist circumference, HDL cholesterol, LDL cholesterol, triglycerides, HbA1c, diastolic blood pressure, and systolic blood pressure. The score was computed when data were available for at least six of the eight biomarkers, allowing for up to two missing values per participant. To ensure consistency in directionality— where higher scores indicate poorer cardiometabolic health— the standardized HDL cholesterol values were inverted prior to averaging.

### Covariates

Covariates were selected as confounders in the analyses a priori based on data availability and known associations with sleep and cardiometabolic health markers. The following covariates were collected in all cohorts: age (years), sex (male, female), device-measured moderate-to-vigorous physical activity (including running, cycling, inclined stepping, and walking with cadence ≥100 steps/min, min/day), smoking status (non-smoker, current smoker), alcohol consumption (tertiles based on self-reported weekly consumption), lipid-modifying, antihypertensive or glucose-lowering medications (yes, no), and history of cardiovascular disease (yes, no). Additionally, a subset of cohorts had data on education (four cohorts; ranging from none or lower than high school to university degree or higher), self-rated health (five-point Likert scale), self-reported diet quality (low, medium, or high vegetable and fruit consumption) and physical function score; which were adjusted for as part of sensitivity analysis. Full details of the measurement and subsequent harmonization of covariates in each cohort are provided in **Supplemental Table 4**.

### Statistical analyses

The range of all cardiometabolic markers and three sleep indices were Winsorized at the 2.5^th^ and 97.5^th^ percentile to alleviate the impact of sparse data or outliers. We conducted a pooled individual participant data analysis using adjusted generalized linear regression to examine the associations between three categorical sleep exposures and composite cardiometabolic health, as well as individual markers including BMI, waist circumference, HDL cholesterol, LDL cholesterol, triglycerides, HbA1c, and both diastolic and systolic blood pressure. Results are presented as visualizations of beta coefficients (β) with 95% confidence intervals (CI). Additionally, we performed joint association analyses to assess all combinations of sleep duration, regularity, and efficiency consisting of 9 mutually exclusive categories for each combination. Further, we examined a three-way interaction analysis of all sleep parameters (27 combinations) with cardiometabolic health markers, utilizing adjusted generalized linear regression models. Regression assumptions were evaluated through residual diagnostics, including residual plots and leverage versus residual squared plots. All models were adjusted for age, sex, cohort, smoking, alcohol consumption, medication use, previous cardiovascular incidence and moderate-to-vigorous physical activity; and mutually adjusted for sleep duration or regularity or efficiency, as appropriate.

To assess the associations of sleep exposures as continuous variables, we employed restricted cubic spline adjusted generalized linear models with knots placed at the 10^th^, 50^th^, and 90^th^ percentiles. Non-linearity was examined using Wald tests. Sensitivity analyses were conducted to assess the robustness of findings by further adjusting for diet, education, self-reported health, and physical function in the three-way joint models from available participants. To reduce the influence of reverse causation, analyses were repeated after excluding participants with self-reported health as poor or fair (excluded n=1,572), excluding participants with prevalent or history of cardiovascular disease (excluded n=1,448), and excluding people with a BMI over 35 kg/m^2^ (excluded n=716). We added a sensitivity analysis with an alternative definition of composite cardiometabolic risk score that uses the same set of eight biomarkers on complete cases, but applies a stricter criterion by excluding individuals with missing data in any biomarkers (excluded n=2,380). We further conducted a sensitivity analysis in which we included only those with at least 7 days of valid wear days (≥20 hours per day) (excluded n=2,976). Interaction effects were tested between each sleep exposure, age, and sex. All statistical analyses were performed using R (version 4.4.0), with modelling conducted using the rms package (version 6.8.1).

## Results

The analytical sample comprised 14,085 participants, with a mean age of 54.3 years and 54.9% were female. Average sleep duration in the pooled cohort was 7.2 (SD: 1.0) hours, with a mean sleep regularity index of 78.4% and a sleep efficiency of 86.8%. **Table 1** shows the baseline characteristics of participants stratified by sleep duration. Compared to those with adequate sleep duration (7–8 hours), short sleepers (<7 hours) were older, had a lower proportion of females, higher smoking rates, poorer self-rated health, greater medication use, a higher prevalence of cardiovascular disease, and higher levels of MVPA. Short sleepers also had a lower level of education and more often had a lower occupational class. Long sleepers (>8 hours) had a higher proportion of females, reported poorer self-rated health, and had lower physical activity levels. The baseline characteristics stratified by SRI and sleep efficiency are shown in **Supplemental Table 5 and 6**. The correlation between sleep duration and both sleep efficiency and sleep regularity was low (r = 0.28 and r = – 0.16, respectively). Sleep efficiency showed a moderate positive correlation with sleep regularity (r = 0.45).

**Table 1.** Baseline characteristics of participants stratified by sleep duration.

| Baseline characteristic | Short<br>($< 7$<br>hours) | Adequate<br>(7-8<br>hours) | Long<br>($> 9$<br>hours) | Full<br>sample |
| --- | --- | --- | --- | --- |
| Sample size | 6,008 | 5,179 | 2,898 | 14,085 |
| Age (mean (SD)) | 55.3<br>(9.5) | 53.7<br>(9.4) | 53.2<br>(9.4) | 54.3<br>(9.5) |
| Sex = Female, n (%) | 2,714<br>(45.2) | 3,064<br>(59.2) | 1,954<br>(67.4) | 7,732<br>(54.9) |
| Cohort, n (%) |  |  |  |  |
| ALSWH | 250<br>(4.2) | 384<br>(7.4) | 284<br>(9.8) | 918<br>(6.5) |
| BCS70 | 1,769<br>(29.4) | 1,822<br>(35.2) | 1,195<br>(41.2) | 4,786<br>(34.0) |
| DPhacto | 231<br>(3.8) | 158<br>(3.1) | 37 (1.3) | 426<br>(3.0) |
| FIREA | 91<br>(1.5) | 45 (0.9) | 11 (0.4) | 147<br>(1.0) |
| NES | 269<br>(4.5) | 201<br>(3.9) | 65 (2.2) | 535<br>(3.8) |
| TMS | 3,398<br>(56.6) | 2,569<br>(49.6) | 1,306<br>(45.1) | 7,273<br>(51.6) |
| **Alcohol consumption<sup>7</sup>, n (%)** |  |  |  |  |
| Tertile 1 (least) | 1,667<br>(31.6) | 1,529<br>(34.0) | 951<br>(39.2) | 4,147<br>(34.0) |
| Tertile 2 (moderate) | 1,778<br>(33.7) | 1,559<br>(34.7) | 808<br>(33.3) | 4,145<br>(34.0) |
| Tertile 3 (highest) | 1,829<br>(34.7) | 1,407<br>(31.3) | 665<br>(27.4) | 3,901<br>(32.0) |
| Smoking = Current smoker, n (%) | 932<br>(15.6) | 651<br>(12.6) | 392<br>(13.6) | 1,975<br>(14.1) |
| **Education, n (%)** |  |  |  |  |
| None or less than high school | 595<br>(10.6) | 518<br>(10.5) | 412<br>(14.6) | 1,525<br>(11.4) |
| High school ($\geq 16$ years) | 1,500<br>(26.7) | 1,313<br>(26.7) | 870<br>(30.9) | 3,683<br>(27.6) |
| Further education ($\geq 16$–18 years) | 2,338<br>(41.6) | 1,929<br>(39.2) | 937<br>(33.3) | 5,204<br>(39.0) |
| University degree or higher | 1,182<br>(21.1) | 1,157<br>(23.5) | 595<br>(21.1) | 2,934<br>(22.0) |
| **Self-rated health, n (%)** |  |  |  |  |
| Excellent | 666<br>(11.3) | 665<br>(13.0) | 335<br>(11.8) | 1,666<br>(12.0) |
| Very good | 1,826<br>(31.0) | 1,733<br>(34.0) | 870<br>(30.5) | 4,429<br>(32.0) |
| Good | 2,648<br>(44.9) | 2,127<br>(41.7) | 1,181<br>(41.5) | 5,956<br>(43.0) |
| Fair | 661<br>(11.2) | 507<br>(9.9) | 365<br>(12.8) | 1,533<br>(11.1) |
| Poor | 93<br>(1.6) | 65 (1.3) | 98 (3.4) | 256<br>(1.8) |
| **Cardiometabolic medication use<sup>1</sup>, n (%)** | 1,973<br>(35.0) | 1,319<br>(26.9) | 805<br>(28.5) | 4,097<br>(30.7) |
| **Prevalent CVD, n (%)** | 660<br>(11.1) | 452<br>(8.8) | 292<br>(10.1) | 1,404<br>(10.0) |
| **MVPA mins per day (mean (SD))<sup>5</sup>** | 78.1<br>(29.5) | 74.4<br>(27.6) | 63.3<br>(26.5) | 73.7<br>(28.7) |
| **Diet<sup>6</sup>, n (%)** |  |  |  |  |
| Low quality | 1,393<br>(38.5) | 1,135<br>(38.8) | 691<br>(45.3) | 3,219<br>(39.9) |
| Medium quality | 1,370<br>(37.9) | 1,134<br>(38.7) | 569<br>(37.3) | 3,073<br>(38.1) |
| High quality | 852<br>(23.6) | 659<br>(22.5) | 264<br>(17.3) | 1,775<br>(22.0) |
| **Body Mass Index, kg/m<sup>2</sup> (mean (SD))** | 27.2<br>(4.5) | 26.4<br>(4.3) | 27.0<br>(4.6) | 26.8<br>(4.5) |
| **Waist circumference, cm (mean (SD))** | 95.7<br>(13.2) | 92.0<br>(12.8) | 93.1<br>(13.3) | 93.8<br>(13.2) |
| **HDL<sup>2</sup>, mmol/L (mean (SD))** | 1.5<br>(0.4) | 1.6 (0.4) | 1.6 (0.4) | 1.6 (0.4) |
| **LDL<sup>3</sup>, mmol/L (mean (SD))** | 3.4<br>(0.9) | 3.4 (0.9) | 3.4 (0.9) | 3.4 (0.9) |
| **Triglycerides, mmol/L (mean (SD))** | 1.5<br>(0.8) | 1.4 (0.7) | 1.5 (0.8) | 1.4 (0.8) |
| **HbA1C<sup>4</sup>, mmol/mol (mean (SD))** | 38.4<br>(7.4) | 37.0<br>(6.5) | 37.5<br>(6.9) | 37.7<br>(7.0) |
| **Systolic blood pressure, mmHg (mean (SD))** | 131.7<br>(16.6) | 128.7<br>(16.4) | 127.6<br>(16.6) | 129.8<br>(16.6) |
| **Diastolic blood pressure, mmHg (mean (SD))** | 77.1<br>(9.9) | 76.0<br>(9.8) | 75.9<br>(9.8) | 76.5<br>(9.8) |
| **Sleep regularity index (mean (SD))** | 79.4<br>(10.1) | 79.6<br>(9.6) | 73.9<br>(11.9) | 78.4<br>(10.6) |
| **Sleep duration, h/day (mean (SD))** | 6.2<br>(0.6) | 7.5 (0.3) | 8.6 (0.5) | 7.2 (1.0) |
| **Sleep efficiency (mean (SD))** | 84.4<br>(9.7) | 89.1<br>(7.1) | 87.7<br>(8.0) | 86.8<br>(8.7) |

SRI: Sleep Regularity Index; SD: standard deviation.
<sup>1</sup>Lipid-modifying, hypertensive, and glucose-lowering medications; <sup>2</sup>High-density lipoprotein cholesterol;
<sup>3</sup>Low-density lipoprotein cholesterol, <sup>4</sup>Glycated Hemoglobin, <sup>5</sup>Accelerometer-derived moderate-to-vigorous physical activity, <sup>6</sup> Self-reported daily intake of fruits and vegetables served as a proxy for dietary, <sup>7</sup>Self-reported alcohol consumption.

### Sleep duration and cardiometabolic health

Compared to those with adequate sleep duration, participants with shorter sleep duration had significantly higher composite cardiometabolic risk z-score (β: 0.05, 95%CI: 0.03-0.07) in the fully adjusted model including adjustment for sleep regularity and efficiency (**Figure 1A**). Long sleep duration was also associated with higher composite cardiometabolic risk score (β: 0.01, 95%CI: - 0.03-0.03), albeit not statistically significant. **Supplemental Figure 1** shows the association of sleep duration with the individual cardiometabolic health markers. Compared to an adequate sleep duration, short sleep duration but not long sleep duration was associated with a significantly higher BMI (β: 0.56, 95%CI: 0.39-0.74); waist circumference (β: 1.65, 95%CI: 1.18-2.12); HbA1c (β: 0.58, 95%CI: 0.31-0.85); and diastolic blood pressure (β: 0.43, 95%CI: 0.02-0.84). There was no association between sleep duration and HDL, LDL cholesterol and triglyceride levels.

**Figure 1.**
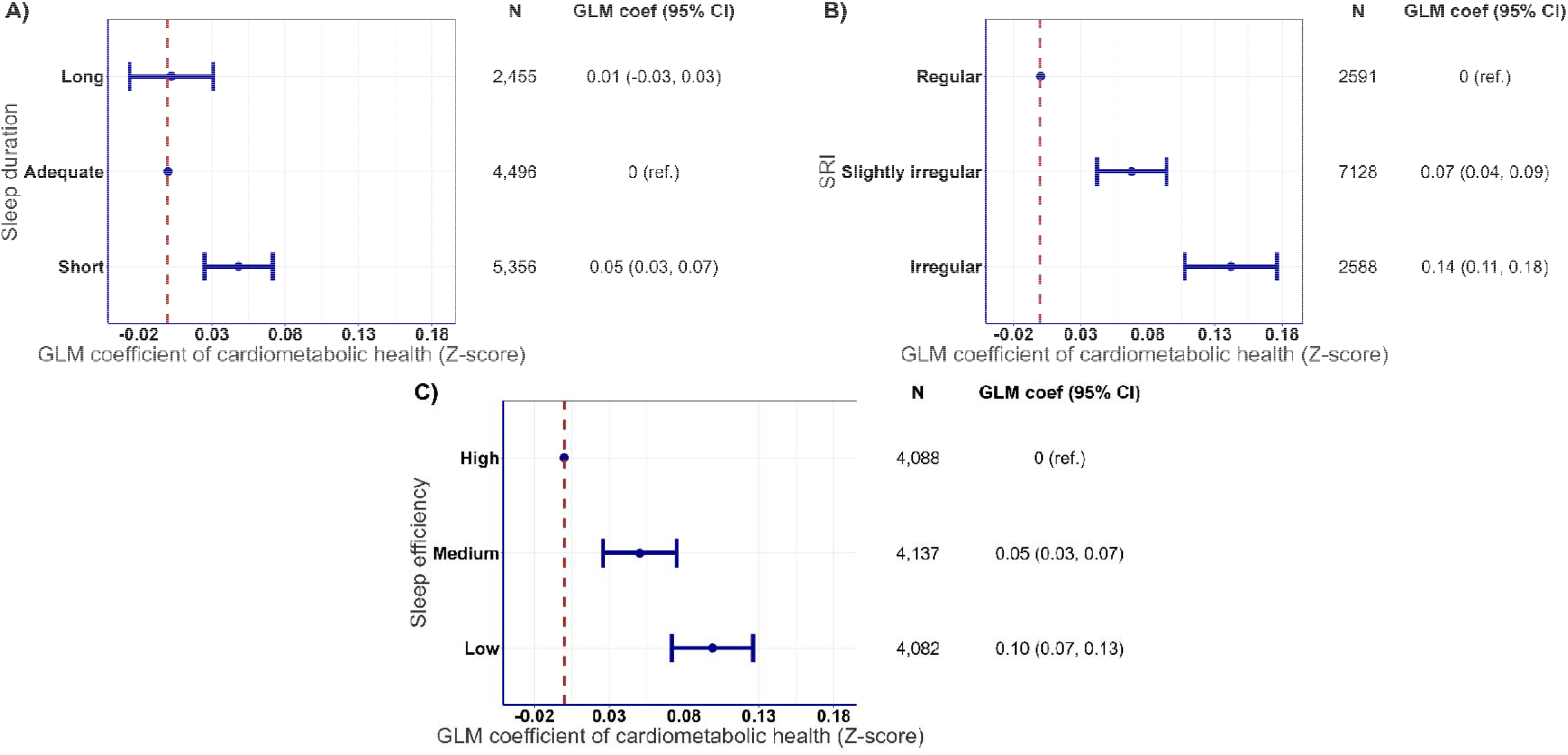
Association of sleep duration (A), regularity (B), and efficiency (C) with composite cardiometabolic risk score. Composite cardiometabolic risk score (z-score) was derived from the composite sample distribution of BMI, waist circumference, HDL cholesterol, LDL cholesterol, triglycerides, glycated haemoglobin (HbA1c), systolic and diastolic blood pressure. A higher score represents poorer cardiometabolic health. Sleep duration was categorized into short (<7h/day), adequate (7-8h/day) and long (>8h/day), sleep regularity (irregular, SRI<71.6; slightly irregular, 71.6≤SRI≤ 87.3; regular, SRI> 87.3) and sleep efficiency (low, efficiency<85.3; medium, 85.3≤ efficiency ≤ 91.8; high, efficiency >91.8) were categorized into tertiles. Reference group was set to adequate, regular and high for sleep duration, SRI and efficiency. All models were adjusted for age, sex, cohort, smoking, alcohol consumption, medication use, previous cardiovascular incidence and moderate-to-vigorous physical activity. All sleep characteristics were mutually adjusted. GLM coefficients represent the mean differences between the reference group and each of the other groups. N=12,307. SRI: Sleep regularity index, BMI: body mass index.

### Sleep regularity and cardiometabolic health

Participants who had a slightly irregular or irregular sleeping pattern had a higher composite cardiometabolic risk z-score compared to those with regular SRI in the fully adjusted model which included adjustment for sleep duration and efficiency (slightly irregular: β: 0.07, 95%CI; 0.04-0.09; irregular: β: 0.14, 95%CI: 0.11-0.18 **Figure 1B**). As for the individual cardiometabolic health markers (**Supplemental Figure 2**), compared to a regular sleep pattern, both slightly irregular and irregular sleep patterns were associated with a higher BMI, a larger waist circumference, lower HDL levels, higher LDL, higher levels of triglycerides, higher HbA1c, and higher diastolic blood pressure. There was no association with systolic blood pressure.

### Sleep efficiency and cardiometabolic health

A lower sleep efficiency was also associated with a higher composite cardiometabolic risk z-score in the fully adjusted model which included adjustment for sleep duration and regularity (**Figure 1C**). Compared to participants with high sleep efficiency, those with a medium and low sleep efficiency had a higher composite cardiometabolic risk score (medium: β: 0.05, 95%CI: 0.03-0.07, low: β: 0.10, 95%CI: 0.07-0.13). As for the individual cardiometabolic health markers (**Supplemental Figure 3**), a medium and low sleep efficiency was associated with a higher BMI, a larger waist circumference, lower HDL levels (only statistically significant for low sleep efficiency), higher levels of triglycerides, higher HbA1c (only statistically significant for low sleep efficiency), and higher systolic and diastolic blood pressure.

### Continuous sleep characteristics and cardiometabolic health

Significant non-linear associations (p<0.05) were observed for sleep duration (**Figure 2A**) and regularity (Figure 2B) but not for sleep efficiency (Figure 2C) with the composite outcome. A reverse J-shaped association was observed for sleep duration and consistent with the categorization above, short sleep duration was associated with a higher composite cardiometabolic risk score with the most favourable cardiometabolic health in those sleeping 7-8 hours. Greater sleep regularity and efficiency were linked to better cardiometabolic profiles.

**Figure 2.**
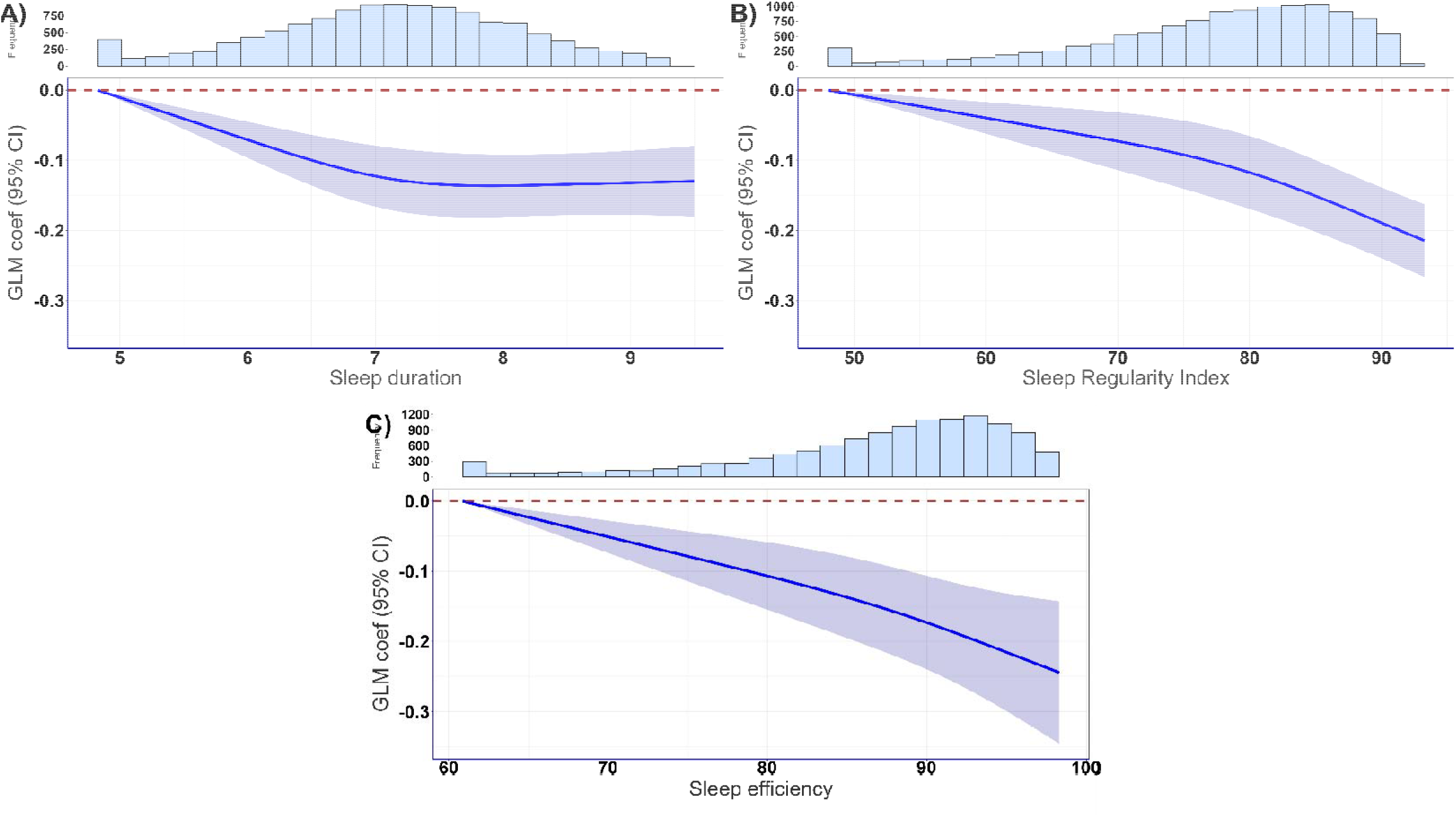
Association of (A) sleep duration, (B) regularity, and (C) efficiency with composite cardiometabolic risk score. Composite cardiometabolic risk score (z-score) was derived from the composite sample distribution of BMI, waist circumferenc HDL cholesterol, LDL cholesterol, triglycerides, glycated haemoglobin (HbA1c), systolic and diastolic blood pressure. A higher score represents poorer cardiometabolic health. Sleep duration, sleep regularity and sleep efficiency were continuous. Reference for each group was the lowest value in the distribution. Knots for each exposure were placed at 10^th^, 50^th^ and 90^th^ percentiles. All models were adjusted for age, sex, cohort, smoking, alcohol consumption, medication use, previous cardiovascular incidence and moderate-to-vigorous physical activity. All sleep characteristics were mutually adjusted. N=12,307. SRI: Sleep regularity index, BMI: body mass index.

### Joint associations of sleep duration, regularity and efficiency with cardiometabolic health

The joint associations of sleep duration and sleep regularity are shown in **Figure 3**. Compared to adequate sleepers with a regular SRI, individuals with more irregular sleep patterns tended to show higher cardiometabolic risk scores in long, adequate, and short sleepers; the highest cardiometabolic risk score was in short sleepers with an irregular sleep pattern (β: 0.19, 95%CI: 0.14-0.25). **Figure 4** shows the joint association between sleep duration and efficiency indicating that compared to individuals with an adequate sleep duration and a high sleep efficiency, those with a medium and low sleep efficiency had a significantly higher cardiometabolic risk score in all sleep duration groups. Finally, **Figure 5** shows joint associations of sleep regularity and sleep efficiency. Compared to those with regular and highly efficient sleep, worse cardiometabolic health was observed in those with a more irregular sleep pattern in those with a medium and low sleep efficiency. The highest cardiometabolic risk score was observed in people with an irregular sleep pattern and a low sleep efficiency (β: 0.24, 95%CI:0.20-0.28).

**Figure 3.**
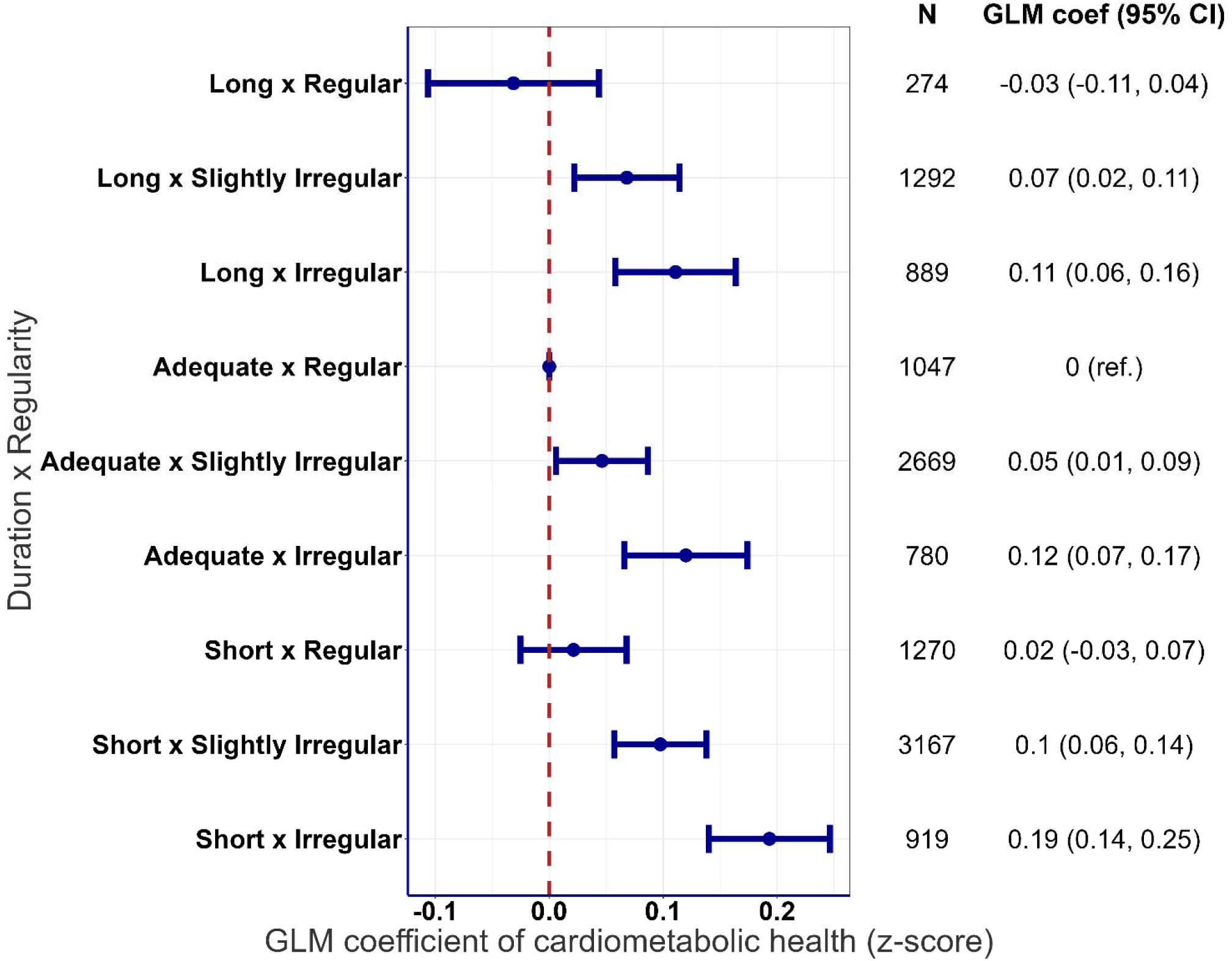
Joint association of sleep duration and regularity with composite cardiometabolic risk score. Sleep duration was categorized into short (<7h/day), adequate (7-8h/day) and long (>8h/day) and sleep regularity (irregular, SRI<71.6; slightly irregular, 71.6≤SRI≤ 87.3; regular, SRI> 87.3) were categorized into tertiles. Reference group was set to adequate duration with regular sleep. All models were adjusted for age, sex, cohort, smoking, alcohol consumption, medication use, previous cardiovascular incidence, moderate-to-vigorous physical activity and sleep efficiency. N=12,307. GLM coefficients represent the mean differences between the reference group and each of the other groups. SRI: sleep regularity index.

**Figure 4.**
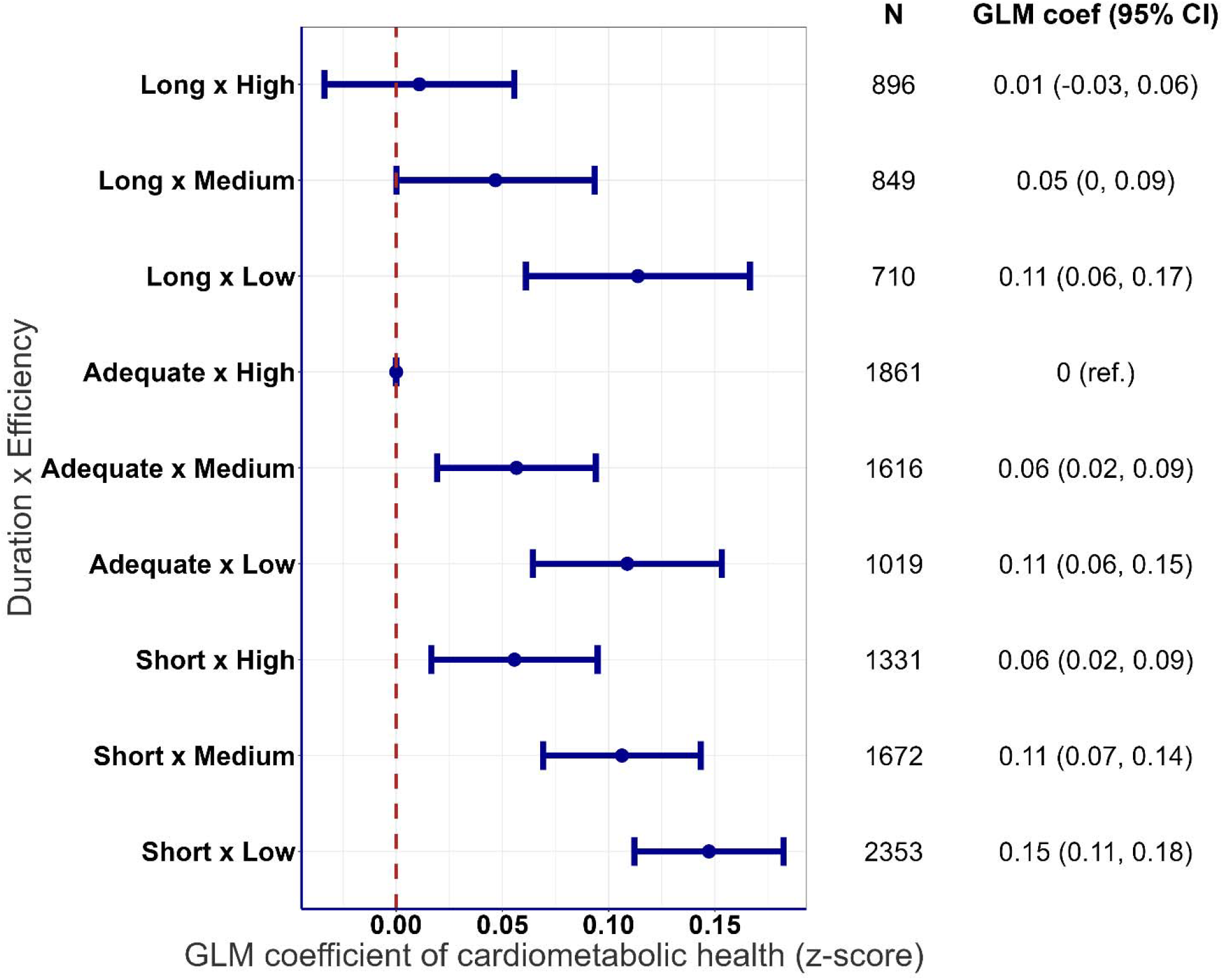
Joint association of sleep duration and efficiency with composite cardiometabolic risk score. Sleep duration was categorized into short (<7h/day), adequate (7-8h/day) and long (>8h/day) and sleep efficiency (low, efficiency<85.3; medium, 85.3≤ efficiency ≤ 91.8; high, efficiency >91.8) were categorized into tertiles. Reference group was set to adequate duration with high efficiency of sleep. All models were adjusted for age, sex, cohort, smoking, alcohol consumption, medication use, previous cardiovascular incidence, moderate-to-vigorous physical activity and sleep regularity. N=12,307. GLM coefficients represent the mean differences between the reference group and each of the other groups. SRI: sleep regularity index.

**Figure 5.**
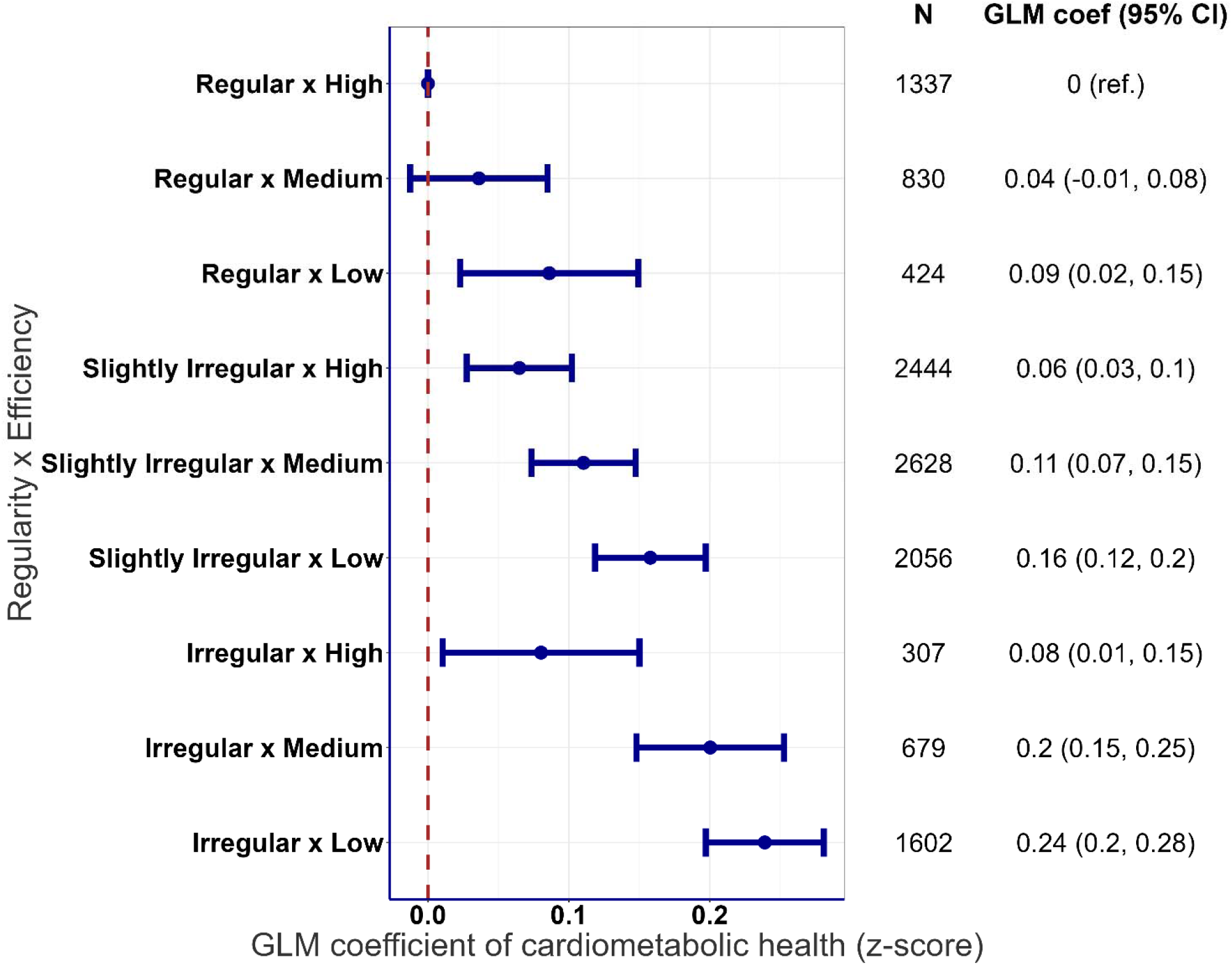
Joint association of sleep regularity and efficiency with composite cardiometabolic risk score. Sleep regularity (irregular, SRI<71.6; slightly irregular, 71.6≤SRI≤ 87.3; regular, SRI> 87.3) and sleep efficiency (low, efficiency<85.3; medium, 85.3≤ efficiency ≤ 91.8; high, efficiency >91.8) were categorized into tertiles. Reference group was set to regular with high efficiency of sleep. All models were adjusted for age, sex, cohort, smoking, alcohol consumption, medication use, previous cardiovascular incidence, moderate-to-vigorous physical activity and sleep duration. N=12,307. GLM coefficients represent the mean differences between the reference group and each of the other groups. SRI: sleep regularity index.

The three-way joint associations of sleep characteristics are presented in **Figure 6**. Compared to the group with an adequate sleep duration, regular sleep pattern, and high sleep efficiency, cardiometabolic risk score was highest in groups with medium or low sleep efficiency, combined with an irregular sleep pattern in people with an adequate and long sleep duration. In the short sleepers, all combinations of sleep regularity and efficiency presented worse cardiometabolic health except for the group with a regular SRI and a high sleep efficiency.

**Figure 6.**
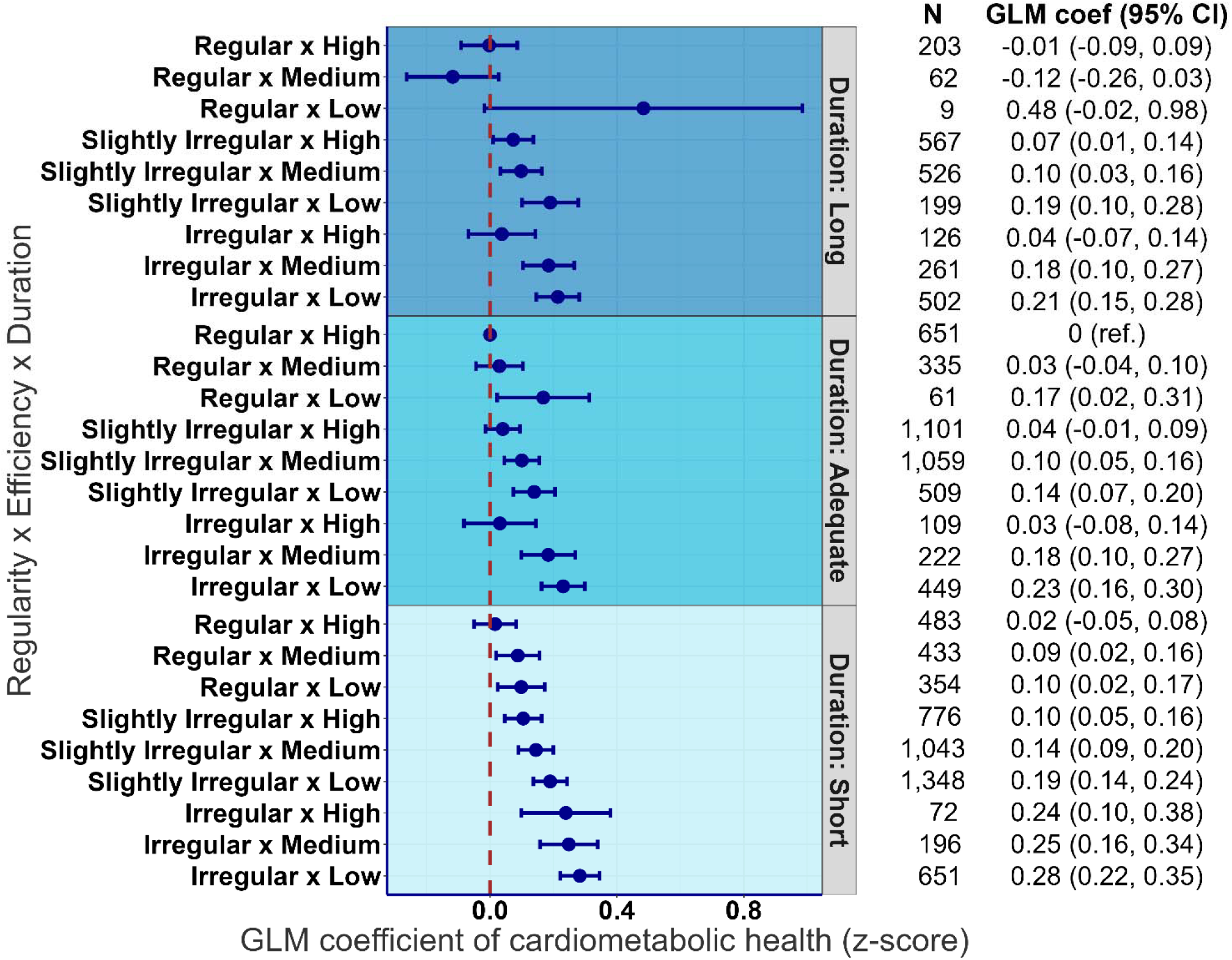
Three-way joint association of sleep duration, regularity and efficiency with composite cardiometabolic risk score. Sleep duration was categorized into short (<7h/day), adequate (7-8h/day) and long (>8h/day), sleep regularity (irregular, SRI<71.6; slightly irregular, 71.6≤SRI≤ 87.3; regular, SRI>87.3) and sleep efficiency (low, efficiency<85.3; medium, 85.3≤ efficiency ≤ 91.8; high, efficiency >91.8) were categorized into tertiles. Reference group was set to adequate duration with regular and high efficiency of sleep. All models were adjusted for age, sex, cohort, smoking, alcohol consumption, medication use, previous cardiovascular incidence and moderate-to-vigorous physical activity. N=12,307. GLM coefficients represent the mean differences between the reference group and each of the other groups. SRI: sleep regularity index.

### Sensitivity analyses

Adjustment for diet and levels of education or physical function did not materially change the results of the joint associations of sleep duration, regularity and efficiency with cardiometabolic risk score (**Supplemental Figure 4** and **Supplemental Figure 6**). Additional adjustment for body fat percentage, which was only available in two of the six cohorts (n=3,373), reduced the regression coefficients; the combination of a long sleep duration with a slightly/irregular patterns and a medium/low sleep efficiency retained significantly higher composite cardiometabolic risk z-scores compared to those with an adequate sleep duration, a regular SRI and a high sleep efficiency (**Supplemental Figure 5**). Results remained similar where participants with poor or fair self-reported health were excluded (**Supplemental Figure 7**); in those with a history of cardiovascular disease were excluded (**Supplemental Figure 8**); in participants with a BMI>35 kg/m^2^ excluded (**Supplemental Figure 9**); with the application of an alternative composite cardiometabolic risk score (**Supplemental Figure 10**); or in participants with at least 7 days of valid wear days (**Supplemental Figure 11**). Multiplicative interactions between each sleep parameter and sex and age were tested; however, no consistent evidence of effect modification by sex or age was observed (**Supplemental Table 7** and **Supplemental Table 8**).

## Discussion

In this individual participant data pooled analysis of six cohort studies within the ProPASS consortium, we investigated the independent and joint associations of device-measured sleep duration, regularity, and efficiency with cardiometabolic health outcomes. In these cross-sectional analyses, shorter sleep duration, greater irregularity in sleep patterns, and lower sleep efficiency were independently associated with poorer cardiometabolic health outcomes compared to adequate sleep duration, regular sleep patterns, and high sleep efficiency respectively. Both the 2-way and 3-way joint associations of sleep duration with sleep regularity and sleep efficiency indicated that, regardless of sleep duration, both greater sleep irregularity and lower sleep efficiency were linked to adverse cardiometabolic health profiles. These associations were particularly pronounced in the 3-way joint analysis, suggesting possible additive effects of sleep efficiency and regularity; individuals with adequate, long, or short sleep duration had worse cardiometabolic health when they also exhibited either a more irregular sleep pattern or lower sleep efficiency. To our knowledge, our study is the first to investigate the 3-way joint associations of any sleep parameters with cardiometabolic health. Our finding that irrespective of sleep duration, both sleep regularity and sleep efficiency are associated with cardiometabolic health is novel, suggesting the importance of sleep domains other than duration for better cardiometabolic health.

Our study suggests that, compared to an adequate sleep duration, short sleep duration—but not long sleep duration—was associated with poorer cardiometabolic health. This association was consistently observed across individual cardiometabolic health markers, except for elevated triglyceride levels in long sleepers, as well as the composite cardiometabolic risk score. This is consistent with findings from a systematic review and meta-analyses showing that short sleep duration was associated with increased risk of mortality, diabetes, hypertension, cardiovascular disease, and obesity.^4^ Our findings that long sleep duration was not associated with cardiometabolic health contrast with previous evidence from a review, which reported that long sleep duration is associated with increased incidence of mortality and several conditions, including stroke, diabetes, and cardiovascular disease, although no significant association was found with hypertension.^3^ An umbrella review synthesizing evidence from 11 systematic reviews - 96% of the underlying studies relied on self-reported sleep data - found a U-shaped association between sleep duration and health outcomes.^1^ The dose–response curves from that review align with the findings of the current study indicating that the most favourable sleep duration was approximately 7–8 hours per day which we chose as the reference group in our analyses. It is important to highlight that the evidence underlying the aforementioned reviews primarily relies on self-reported sleep duration. This is noteworthy because long sleep duration is often overestimated in self-reports, and a recent study has shown that self-reported—but not objectively measured—sleep duration was associated with all-cause and cardiovascular disease mortality.^36^

Our results provide novel empirical evidence suggesting that an irregular sleep pattern and low sleep efficiency are both associated with poorer cardiometabolic health, independent of each other and sleep duration. Meta-analyses and systematic reviews have shown that poor sleep quality - as assessed by self-reported measures in the majority of studies - was associated with coronary heart disease,^5^ hypertension^37^, arterial stiffness^38, 39^ and endothelial dysfunction^39^ but not with all-cause or rular disease mortality^5^ or atherosclerosis.^39^ Systematic reviews on sleep variability - based on self-reported data or accelerometry - and cardiometabolic health have shown associations with obesity, ^9, 40^ cardiovascular disease, and metabolic syndrome^9,41^ but not with insulin resistance.^9^ It is important to note that sleep variability in the mostly observational studies underlying these reviews has been assessed with different indicators such as ‘social jetlag’ as the variation between working days and free days, and the standard deviation of sleep duration of sleep across multiple days. We chose to operationalize the SRI as this parameter is increasingly prioritized as an objective metric for day-to-day variability in sleep wake cycle. A recent study using data from the UK Biobank investigating joint associations between sleep duration and sleep regularity shows that meeting sleep duration recommendations did not offset the adverse effect of an irregular sleep patterns for the risk type 2 diabetes and major adverse cardiovascular events.^8 14^

In a 2023 consensus statement by the US National Sleep Foundation, the expert panel acknowledged the importance of sleep regularity for health and performance.^41^ Further, a recent 2025 scientific statement from the American Heart Association outlines the concept of multidimensional sleep health and highlights the multiple dimensions of sleep that are relevant for cardiometabolic health.^42^ It emphasizes that while sleep duration has been widely studied and thereby informed the development of guidelines, more research is needed to examine multiple sleep dimensions. It is important to note that the American Heart Association’s construct for cardiovascular health ‘Life’s Essential 8’ includes sleep duration as a key factor for cardiovascular health.^43^ This broader focus on multiple dimension of sleep is necessary to support the creation of comprehensive public health guidelines and clinical interventions.

While there is substantial evidence to support the biological causal link between short sleep duration and adverse health outcomes, the biological pathway of long sleep and health remains less clear. Long sleep may be related to poorer sleep quality and sleep fragmentation imposed by other health conditions; therefore there is a possibility that it reflects reverse causation or residual confounding more than causal effects on cardiometabolic health.^44^ In the current study, our findings indicate that individuals with long sleep duration combined with either an irregular sleep pattern or low sleep efficiency exhibit poorer cardiometabolic health. This finding suggests that it is not long sleep duration, but its interaction with other adverse sleep characteristics that is associated with negative health outcomes. Sleep irregularity may result in circadian misalignment, a disconnect between the central and peripheral clocks. Short-term circadian misalignment has been associated with acute metabolic disturbances including an increase in postprandial glucose, insulin and mean arterial pressure, decreases in leptin and sleep efficiency,^45^ increases in 24-hour blood pressure, and inflammation.^10^ These mechanisms may explain how circadian misalignment may lead to increased cardiovascular disease risk.^9^

Strengths of our study include the large sample size (n=14,085) drawn from six cohorts across five countries and the device-based assessment of multiple sleep parameters. By examining the joint associations of three sleep characteristics with cardiometabolic health, our study contributes to the growing body of evidence that sleep is a multidimensional construct, which should be evaluated beyond sleep duration alone. Our study also has some limitations that should be acknowledged. First, our study was cross-sectional, and reverse causation is possible - that is, poorer health may have led to less optimal sleep parameters. A limitation of relying on accelerometry alone to estimate sleep parameters is that it is less accurate in distinguishing between quiet reclined wakefulness and actual sleep^17,18^, potentially leading to exposure misclassification and underestimation of any true associations that may exist. Overall, validation work has shown that a thigh-worn accelerometer can measure sleep duration with comparable accuracy as a wrist-worn accelerometer. ^18^ Further, we did not have data on clinical sleep disorders such as sleep apnoea or insomnia. Finally, adjustments were limited to selected confounders that were available in all cohorts and we cannot exclude residual confounding. Some of the additional adjustments, e.g. for shift work was not possible because of data availability and additional adjustment for fat mass was only possible in a smaller sample as it was not measured in all cohorts.

## Conclusion

These findings suggest the importance of adopting a multidimensional approach to sleep health, demonstrating that sleep duration alone may not fully capture its relationship with cardiometabolic risk. By demonstrating the joint associations of sleep duration, regularity, and efficiency, this study contributes to the growing evidence that multidimensional sleep profiles are more informative than single sleep parameters such as duration. Future longitudinal studies are needed to further clarify these associations across diverse populations and to evaluate the effectiveness of interventions aimed at promoting consistent sleep patterns and enhancing sleep efficiency as strategies to reduce cardiometabolic risk.

## Data Availability

All data produced in the present study are available upon reasonable request to the authors

## Acknowledgements

The data on which this research is based were drawn from six observational studies in the Netherlands, UK, Australia, Denmark, and Finland. We are grateful to all participants who provided the survey data.

## Details of individual cohort funding

The Maastricht Study was supported by the European Regional Development Fund via OP-Zuid, the Province of Limburg, the Dutch Ministry of Economic Affairs (grant 31O.041), Stichting De Weijerhorst (Maastricht, the Netherlands), the Pearl String Initiative Diabetes (Amsterdam, the Netherlands), the Cardiovascular Center (CVC, Maastricht, the Netherlands), CARIM School for Cardiovascular Diseases (Maastricht, the Netherlands), CAPHRI Care and Public Health Research Institute (Maastricht, the Netherlands), NUTRIM School for Nutrition and Translational Research in Metabolism (Maastricht, the Netherlands), Stichting Annadal (Maastricht, the Netherlands), Health Foundation Limburg (Maastricht, the Netherlands), and by unrestricted grants from Janssen-Cilag BV (Tilburg, the Netherlands), Novo Nordisk Farma BV (Alphen aan den Rijn, the Netherlands), and Sanofi-Aventis Netherlands BV (Gouda, the Netherlands).

The 1970 British Cohort Study is funded by the Economic and Social Research Council (ES/M001660/1). The age 46 sweep was also funded by the British Heart Foundation (grant SP/15/6/31397) and a joint award from the Economic and Social Research Council and the Medical Research Council (grant RES-579-47-0001).

The ALSWH is funded by the Australian Government Department of Health, Disability and Ageing and its substudy (Menarche-to-PreMenopause), from which accelerometry and clinical data were obtained, was funded by the National Health and Medical Research Council Project Grant (APP1129592); and partly supported by NHMRC Investigator (APP1194510) and Ideas (APP1180812) Grants.

DPhacto is funded by The Danish Working Environment Research Fund. NES is supported by the Department of Physiology of the Radboud University Medical Center, the Dutch Heart Foundation (2020T063), and Siemens Healthcare Diagnostics (the Hague, Netherlands).

FIREA is supported by the Academy of Finland (286294, 294154, 319246, 332030, 361780), Ministry of Education and Culture, Juho Vainio Foundation and Finnish State Grants for Clinical Research.

