## Supplemental material for "Associations of Device-Measured Sleep Duration, Regularity, and Efficiency with Cardiometabolic Health in Adults: Findings from the ProPASS Consortium"

**Supplemental Table 1.** Overview of individual cohort details.

**Supplemental Table 2.** Description of sleep variables, relevant references, and source code.

**Supplemental Table 3**. Assessment of cardiometabolic markers across cohort.

**Supplemental Table 4.** Assessment and harmonisation of covariates across cohort.

**Supplemental Table 5**. Baseline characteristics of participants stratified by sleep regularity index (SRI) group.

**Supplemental Table 6**. Baseline characteristics of participants stratified by sleep efficiency.

**Supplemental Table 7**. Interaction between sleep parameters and sex.

**Supplemental Table 8**. Interaction between sleep parameters and age.

**Supplemental Figure 1**. Association of sleep duration with cardiometabolic health markers including A) BMI, B) waist circumstance, C) HDL cholesterol, D) LDL cholesterol, E) Triglycerides, F) Glycated haemoglobin (HbA1c), G) Systolic and H) Diastolic blood pressure.

**Supplemental Figure 2**. Association of sleep regularity with cardiometabolic health markers including A) BMI, B) waist circumstance, C) HDL cholesterol, D) LDL cholesterol, E) Triglycerides, F) Glycated haemoglobin (HbA1c), G) Systolic and H) Diastolic blood pressure.

**Supplemental Figure 3**. Association of sleep efficiency with cardiometabolic health markers including A) BMI, B) waist circumstance, C) HDL cholesterol, D) LDL cholesterol, E) Triglycerides, F) Glycated haemoglobin (HbA1c), G) Systolic and H) Diastolic blood pressure.

**Supplemental Figure 4**. Joint association of sleep duration, regularity and efficiency with composite cardiometabolic risk score with additional adjustment for diet and education.

**Supplemental Figure 5**. Joint association of sleep duration, regularity and efficiency with composite cardiometabolic risk score with additional adjustment for body fat percentage.

**Supplemental Figure 6**. Joint association of sleep duration, regularity and efficiency with composite cardiometabolic risk score with additional adjustment for physical function.

**Supplemental Figure 7**. Joint association of sleep duration, regularity and efficiency with composite cardiometabolic risk score excluding those self-reported health as poor or fair.

**Supplemental Figure 8**. Joint association of sleep duration, regularity and efficiency with composite cardiometabolic risk score excluding those with previous history of cardiovascular disease.

**Supplemental Figure 9**. Joint association of sleep duration, regularity and efficiency with composite cardiometabolic risk score excluding those with BMI over 35.

**Supplemental Figure 10.** Three-way joint association of sleep duration, regularity and efficiency with composite cardiometabolic risk score with an alternative composite cardiometabolic risk score.

**Supplemental Figure 11.** Three-way joint association of sleep duration, regularity and efficiency with at least 7 days of valid wear days (≥20 hours per day).

**Supplemental Table 1.** Overview of individual cohort details.

| **Cohort** | **Sample description and age range** | **Sample size with valid accel data** | **Sex** | **Device** | **Leading institution, Country** | **Eligibility criteria** | **Ethics approval (committee name, reference number)** |
| --- | --- | --- | --- | --- | --- | --- | --- |
| Australian Longitudinal Study on Women’s Health (ALSWH) | General population / 41-49 years | n=941 with accelerometer data | Females only | ActivPAL3 and ActivPAL4 micro | The University of Queensland and The University of Sydney, Australia | Women aged 40-45, not pregnant, not currently undergoing treatment for breast or reproductive cancer | Metro South Health and Health Services Human Research Ethics Committee (reference number: HREC/2019/ QMS/52052) |
| 1970 British Cohort Study (BCS70) | General population /  46 years | n=5229 with accelerometer data | Females & males | ActivPAL3 micro | University College London, United Kingdom | Born within 1 week in April 1970 in England, Scotland or Wales | NRES Committee South East Coast - Brighton and Sussex (Ref 15/LO/1446) |
| Danish PHysical ACTivity cohort with Objective measurements cohort (DPhacto) | Workers in cleaning, manufacturing, and transportation companies /  18-65 years | n=771 with accelerometer data | Females & males | Actigraph GT3X | National Research Centre for the Working Environment, Denmark | Workers from manual-based jobs in manufacturing, transportation and cleaning sectors | Danish data protection agency and local Ethics Committee (H-2-2012-011). |
| Finnish Retirement and Aging Study (FIREA) | General Population, public sector employees / 59-65 years | n=253 with accelerometer data | Females & males | Axivity AX3 | University of Turku, Finland | Public sector employees whose statutory retirement date was between 2014 and 2019 | Ethics Committee of Hospital District of Southwest Finland. |
| Nijmegen Exercise Study (NES) | Participants in Nijmegen 4-day Marches or Seven Hills Run, and their friends & family members /  23-87+ years | n=537 with accelerometer data | Females & males | ActivPAL3 | Radboud university Medical centre, Netherlands | Individuals participating in Dutch sport events (i.e. International Nijmegen Four Days Marches and the Seven Hills Run) and their family and friends. | The Local Ethics Committee on Research Involving Human Subjects (CMO) of the region Arnhem and Nijmegen, the Netherlands (NL36743.091.11) |
| The Maastricht Study (TMS) | General Population (Oversampling of those with T2 Diabetes) /  39-79 years | n=7515 with accelerometer data | Females& males | ActivPAL3 | Maastricht University Medical Center+ and Maastricht University, Netherlands | Individuals aged 40-75 years old, oversampling of those with Type 2 Diabetes | Institutional medical ethical committee (NL31329.068.10) and the Minister of Health, Welfare and Sports of the Netherlands (Permit 131088-105234-PG). |

**Supplemental Table 2** Description of sleep variables, relevant references, and source code.

| **Behaviour** | **How behaviour is derived** | **Key references** | **Source Code** |
| --- | --- | --- | --- |
| Sitting or lying | Using thresholds of inclination of the thigh and movement intensity according to Acti4 algorithm with 1s epoch. | <https://doi.org/10.1123/jpah.2011-0347> | <https://github.com/Ergo-Tools/ActiPASS/blob/main/algorithms/ActivityDetect.m> |
| Differentiation of Sitting and lying | Using the rotation of the thigh each epoch is further differentiated as sitting or lying as described in ActiPASS lying-down algorithm | <https://doi.org/10.3390/s21030904> | <https://github.com/Ergo-Tools/ActiPASS/blob/main/algorithms/lyingAlgB.m> |
| Times-in-bed (primary lying container) | Times-in-bed is derived by considering the periods of lying down (also prolonged-sitting bouts). The two longest consecutive periods of such lying down within a 48-hour moving time window were chosen (with a weighted is to lying periods and periods occurring within 22.00-08.00) as the times-in-bed. | <https://doi.org/10.1123/jpah.2011-0347>  <https://github.com/Ergo-Tools/ActiPASS/wiki/Presentations-about-ActiPASS-usage-and-algorithms#presentation-about-actipass-and-how-it-detects-time-in-bed-primary-lying-container-presented-at-propass-5th-annual-meeting-october-2023> | <https://github.com/Ergo-Tools/ActiPASS/blob/main/algorithms/calcBedLgc.m> |
| Sleep | Each time-in-bed period is further processed by ActiPASS sleep algorithm and each epoch is differentiated as sleep or awake | <https://doi.org/10.1111/jsr.13725> | <https://github.com/Ergo-Tools/ActiPASS/blob/main/algorithms/SkottesSlp.m> |
| Sleep Regularity Index | The Sleep Regularity Index (SRI) quantifies the consistency of an individual’s sleep–wake patterns across consecutive days, based on accelerometer-derived sleep–wake states calculated at the epoch level using an open-source algorithm. It accounts for variability in sleep timing, duration, naps, and awakenings by allowing multiple sleep bouts per day, and is scaled from 0 to 100, representing the probability of being in the same state (sleep or wake) at any two time points 24 hours apart. | <https://doi.org/10.1093/sleep/zsad253> | <https://github.com/dpwindred/sleepreg> |

**Supplemental Table 3**. Assessment of cardiometabolic markers across cohort.

|  | ***Harmonised construct*** | ***Australian Longitudinal Study on Women’s Health*** | ***1970 British Birth Cohort Study*** | ***Danish PHysical ACTivity cohort with Objective measurements cohort*** | ***Finnish Retirement and Aging Study*** | ***Nijmegen Exercise Study*** | ***The Maastricht Study*** |
| --- | --- | --- | --- | --- | --- | --- | --- |
| *Collection of blood samples* |  | Nurses collected non-fasting venous blood samples in morning. | Nurses collected non-venous blood samples at various times throughout day. | n/a | Nurses collected fasting venous blood samples in the morning. | Blood was drawn and freezed (-80 degrees Celsius) and analysed within 5 months. | Trained research staff collected venous blood samples in the morning |
| *Fasted/non-fasted blood sample* | 0: Non-fasted  1: Fasted | 0: All participants provided non-fasted sample | 0: All participants provided non-fasted sample | n/a | 1: All participants provided fasted sample. | 0: Did not fast for 4 hours OR had light meal 4-8 hours before blood draw  1: Fasted for 4 hours AND did not have light meal 4-8 hours before blood-draw | 1: All participants provided fasted sample |
| *Total cholesterol* | Continuous measure (mmol/L) | Measured using routine autoanalyser methods.  QLD/VIC samples  Method principle: Cholesterol oxidase, esterase, peroxidase  Manufacturer: Beckman Coulter DXC800  CV: <1.8%  SA/WA/NSW samples:  Method principle:  Cholesterol oxidase, esterase, peroxidase  Manufacturer: Siemens Atellica  CV: <2% | Method principle: Enzymatic colourimetric- cholesterol esterase/cholesterol oxidase/peroxidase  Manufacturer: Roche Cobas c702, generation 2 assay  CV: ≤2.6% | n/a | Measured with standard (enzymatic and colorimetric) methods    Manufacturer: Cobas 8000 c702, Roche Diagnostics    CV: ≤4.8% | Method principle: Enzymatic- cholesterol esterase and cholesterol oxidase  Manufacturer: Siemens Attelica CH  CV: ≤1.3% | Measured with standard (enzymatic and/or colorimetric) methods  Manufacturer: Beckman Synchron LX20, Beckman Coulter Inc., Brea, USA; or Roche Cobas 6000, Roche diagnostics, Mannheim, Germany  CV: ≤2.5% |
| *SBP and DBP* | Continuous measure (mmHg) | Mean of 2nd and 3rd measurements taken after a 5-min seated rest period using an automated blood pressure monitor (arm not recorded in protocol, although same arm used for all measurements)  Models**:**  QLD: Vital Signs Machine 6000 Series  VIC: Welch Allyn Connex® ProBP™ 3400  SA: Phillips MP30  WA: Omron HEM-907  NSW: Omron HEM-7121 | Mean of 3 measurements on the right arm after a 5-min seated rest period using an Omron HEM 907 blood pressure monitor at 1-minutes intervals. | n/a | Mean of 2 measurements (1 left, 1 right) after a 5-minute seated rest period, using the Microlife Watch BP Office Central monitor. | Mean of 3 measurements (2 left, 1 right) after a 10-min lying rest period using Omron M3 monitor  If differences between measures were >10 mmHg for SBP or >5 mmHg for DBP, a 4th measurement was taken on the right arm and all 4 measurements were used to calculate average. | Mean of 3 measurements on the right arm after a 10-minute seated rest period, using an Omron 705IT monitor.  If differences between 2nd and 3rd measure >10mmHg, a 4th measurement was performed and all 4 measurements were used to calculate average. |
| *HDL* | Continuous measure (mmol/L) | Measured using routine autoanalyser methods.  QLD/VIC samples  Method principle: Direct measure, polymer-polyanion  Manufacturer: Beckman Coulter DXC800  CV: <3%  SA/WA/NSW samples  Method principle:  Direct measure, polymer-polyanion  Manufacturer: Siemens Atellica  CV: <3.2% | Method principle: Enzymatic colourimetric- dextran sulphate/PEG-cholesterol esterase/PEG-cholesterol oxidase/peroxidase.  Manufacturer: Roche Cobas c702, generation 3 assay  CV: ≤2.8% | n/a | Measured with standard (enzymatic and colorimetric) methods    Manufacturer: Cobas 8000 c702, Roche Diagnostics    CV: ≤6.1% | Method priniciple: Enzymatic- cholesterol esterase and cholesterol oxidase  Manufacturer: Siemens Attelica CH  CV: ≤2.0% | Measured with standard (enzymatic and/or colorimetric) methods by an automatic analyzer  Manufacturer: Beckman Synchron LX20, Beckman Coulter Inc., Brea, USA; or Roche Cobas 6000, Roche diagnostics, Mannheim, Germany  CV: ≤4.5% |
| *LDL* | | Continuous measure (mmol/L) | | n/a | LDL was calculated from Friedewald equation (doi: 10.5334/gh.1214) using total cholesterol, HDL and triglycerides. | | |
| *HbA1c* | Continuous measure (mmol/L) | Measured using routine autoanalyser methods.  Method principle: High performance liquid chromatography  Manufacturer: Bio-Rad D100  CV: <1.5% | Method principle: Ion exchange HPLC. Manufacturer: Tosoh G8  CV: ≤3.3% | n/a | n/a | n/a | Method principle: Ion-exchange high performance liquid chromatography  Manufacturer: Variant tm II, Bio-Rad, Hercules, California, USA  CV: ≤1.2% |
| *Triglycerides* | Continuous measure (mmol/L) | Measured using routine autoanalyser methods.  QLD/VIC samples  Method principle: Enzymatic, end point  Manufacturer: Beckman Coulter DXC800  CV: <3.3%  SA/WA/NSW samples  Method principle:  Enzymatic, end point  Manufacturer: Siemens Atellica  CV: <3.4% | Method principle: Enzymatic colourimetric: lipoprotein lipase/glycerol kinase/glycerol phosphate oxidase/peroxidase Manufacturer: Roche Cobas c702  CV: ≤2.4% | n/a | Measured with standard (enzymatic and colorimetric) methods    Manufacturer: Cobas 8000 c702, Roche Diagnostics    CV: ≤5.3% | Method priniciple: Enzymatic- endpoint  Manufacturer: Siemens Attelica CH  CV: ≤2.5% | Measured with standard (enzymatic and/or colorimetric) methods by an automatic analyzer (Beckman Synchron LX20, Beckman Coulter Inc., Brea, USA; or Roche Cobas 6000, Roche diagnostics, Mannheim, Germany  CV: ≤3.5% |
| *BMI* | Continuous measure (kg/m^2^) | Derived from clinical measurement of height (stadiometer) and weight (digital scale)  Stadiometer models:^a^  QLD: ADE (no name)  VIC: Seca BE35208  SA: Seca (no name)  WA: ADE MZ10023  NSW: ADE (no name)  Weigh scale models:^a^  QLD: Seca 813  VIC: Seca BE38844  SA: Soehnle EB9373  WA: Perma Lifestyle Professional Slimline Body Monitor  NSW: Seca 813 | Derived from clinical measurement of height (without shoes; portable Leicester stadiometer  and weight (Tanita BF-522W scales) | Derived from clinical measurement of height (without shoes; stadiometer- Seca, model 213) and weight (Tanita bio-impedance segmental body composition analyzer- model BC418 MA) | Derived from clinical measurement of height (without shoes) and weight  using Inbody 720 scale (Biospace Co.,  Seoul, Korea) | Derived from clinical measurement of height (without shoes) and weight (Seca 881) | Derived from clinical measurement of height (stadiometer – Seca 222) and weight (Seca 877) |
| *Waist circumference* | Continuous measure (cm) | Average of 2 measurements taken with a Seca tape measure to the nearest mm. If measures differed by 5mm, a 3^rd^ measure was taken.  Measured in millimetres at the midpoint between the bottom of the last palpable rib and top of the iliac crest. | Average of 2 measurements taken with a Seca tape measure to nearest mm. If measures differed by 3+ cm, a 3^rd^ measure was taken.  Measured in milimetres midway between the lower rib margin and iliac crest. | Single measurement with an anthropometric tape measure.  Measured in millimetres at a level midway between the lower rib margin and iliac crest | Average of 2 measurements with an anthropometric tape measure and directly on the participant's skin.  Measured in millimetres at a level midway between the lower rib margin and iliac crest. | Single measurement with an anthropometric tape measure directly on the participant’s skin taken when the participant exhaled.  Measured in cm  at a level midway between the lower rib margin and iliac crest. | Average of 2 measurements taken with a flexible plastic tape measure (Seca, Hamburg, Germany).  Measured to the nearest 0.5cm midway between the lower rib margin and the iliac crest at the end of expiration |
| CV: coefficient of variation QLD: Queensland VIC: Victoria SA: South Australia WA: West Australia NSW: New South Wales | | | | | | | |

**Supplemental Table 4.** Assessment and harmonisation of covariates across cohort.

|  | ***Harmonised construct*** | ***Australian Longitudinal Study on Women’s Health*** | ***1970 British Cohort Study*** | ***Danish PHysical ACTivity cohort with Objective measurements cohort*** | ***Finnish Retirement and Aging Study*** | ***Nijmegen Exercise Study*** | ***The Maastricht Study*** |
| --- | --- | --- | --- | --- | --- | --- | --- |
| ***MAIN ANALYSES COVARIATES*** | | | | | | | |
| *Age* | Continuous (years) | Question: “What is your age in years?” | All participants assigned age 46 (birth cohort study; year of birth: 1970; year of accelerometer assessment: 2016) | Determined using workers’ unique civil registration number based on time between date of birth and date of measurement visit | Derived based on time between date of birth and date of measurement visit | Derived based on time between date of birth and date of measurement visit | Derived based on time between date of birth and date of measurement visit |
| *Sex* | 1: Male  2: Female | 2: Female | 1: Male  2: Female | 1: Male  2: Female | 1: Male  2: Female | 1: Male  2: Female | 1: Male  2: Female |
| *Smoking* | 0: Non-smoker  1: Smoker | **Question**: "How often do you currently smoke?"  **Responses & coding**:  0: Not at all  1: Daily; At least weekly (but not daily); Less often than weekly | **Question**: “Which of the statements on this card applies to you?”  **Responses & coding**:  0: I've never smoked cigarettes; I used to smoke but don't at all  1: I now smoke occasionally but not daily; I smoke cigarettes every day | **Question**: “Do you smoke?”  **Responses & coding**:  0: Never smoked; Formerly smoked  1: Daily smoking; Occasionally smoking | **Question**: “Do you currently smoke or have you smoked regularly, i.e. daily or almost daily?”  **Responses & coding**:  0: No I have never smoked; Yes, previously  1: Yes, currently | **Question**: “Do you smoke?”  **Responses & coding**:  0: No, but I smoked in the past; No, I have never smoked  1: Yes | **Question**: “Do you smoke?”  **Responses & coding**:  0: No I have never smoked; No, I stopped smoking more than 6 months ago; No, I stopped less than 6 months ago  1: Yes |
| *Alcohol consumption* | 1: Lowest tertile  2: Middle tertile  3: Highest tertile  (cohort-specific tertiles) | **Question**: “How often do you normally drink alcohol?”  **Responses & coding**:  1: Less than once a week  2: 1-4 days/week  3: 5+ days/week | Units consumed in last 7 days (continuous)  Derived variable from interview questions on type, size and number of alcoholic drinks | **Question**: “Do you drink alcohol? How many units did you drink last week?”  (continuous) | Total intake in grams/week (continuous)  Derived from questionnaire items on amount of beer, wine and spirits | **Question**: “How many glasses did you consumed on average per week in the past year?”  (continuous) | Total intake in grams/day (continuous)  Derived from Food Frequency Questionnaire with a 1-year reference period |
| *Medication use* | 0: No lipid modifying, hypertensive or glucose lowering medications  1: 1 or more of the above | **Protocol:** Participants were asked to bring all medications to the assessment, which were coded using  Anatomical Therapeutic Chemical classification  Anatomical Therapeutic Chemical classification | **Protocol:** Research nurses collected data on all prescription medications which were coded using British National Formulary edition 69 codes  **Responses & coding**:  1: Any of 0212 : Lipid-Regulating Drugs;  0201 – 0207: hypertension related drugs; 0601 Drugs Used In Diabetes | **Questions**: “Have you in the last three months been taken prescription medication?” “If yes, what kind of medication?”  **Responses & coding**:  1: Antihypertensive  0: No medications or other medications  **Questions:** “Do you take medication for high blood pressure?  **Responses & coding**:  1: Yes  2: No | **Protocol:** Research nurses inquired about all prescription medications, which were coded using  Anatomical Therapeutic Chemical classification  1: Any of X01 anti-hypertensive drug;  X02 diabetes drug; X04= cholesterol medicine | **Questions**: “Did you use medication in the past year?” was asked immediately following positive responses to “Which of the following diseases below has been diagnosed by physician?” for 1) hypercholesterolemia; 2) hypertension; 3) diabetes  **Responses & coding**:  1: Yes for any of above  0: No | **Protocol:** Participants were asked to bring all medications to the assessment, which were coded using  Anatomical Therapeutic Chemical classification  Anatomical Therapeutic Chemical classification |
| *History of cardiovascular conditions* | 0: No history of CVD  1: History of CVD | **Questions**:  Wave 1:"Have you ever been told by a doctor that you have heart disease"  Wave 2: "[In the last 4 years] [In more than 4 years ago], have you ever been told by a doctor that you have heart disease?”  Waves 3-8: "In the last 3 years have you been diagnosed or treated for heart disease?"  Wave 8: "In the last 3 years have you been diagnosed or treated for hypertension?"  **Responses & coding**:  0: No to all of above  1: Yes to any of above | **Questions**: “Since last collection wave, have you had any of the health problems listed on this card? [high blood pressure]”  “Since last collection wave, have you had any of the health problems listed on this card? Please include any health problems that had already started before that date. [heart problems]/ [stroke]”.  **Responses & coding**:  0: No to all of above  1: Yes to any of above | **Question**: “Do you have angina pectoris?”  **Responses & coding**:  0: No  1: Yes | **Questions**: “Has a doctor/physician given you a diagnosis of [angina pectoris]/[myocardial infarction]/[stroke]/ [hypertension]?”  **Responses & coding**:  0: No to all of above  1: Yes to any of above | **Questions**: “Which type of diseases below has been diagnosed by physician? [myocardial infarction]/ [heart failure]/ [stroke]/ [atrial fibrillation]/ [hypertension]”  **Responses & coding**:  0: No to all of above  1: Yes to any of above | **Questions**: Rose Questionnaire  Responses:  1: Selected any of myocardial infarction - cerebrovascular infarction and/or hemorrhage - percutaneous artery angioplasty of the coronary arteries, abdominal arteries, peripheral arteries or carotid artery - vascular surgery on coronary arteries, abdominal arteries, peripheral arteries or carotid artery.  0: selected none of the above |
| **SUPPLEMENTARY ANALYSES COVARIATES** | | | | | | | |
| *Education* | **0**: None or lower than high school  **1**: High school qualifications (age 16y)  **2**: Further education qualifications (age 16-18y)  **3**: university degree and higher (18+y) | **Question**: “What is the highest level of qualification you have completed?”  **Responses & coding**:  **0**: No formal qualifications **1**: Year 10 or equivalent  **2**: Year 12 or equivalent, Trade/apprenticeship, Certificate/Diploma  **3**: University degree, Higher university degree | **Derived** **variable** of National Vocational Qualifications categories based on self-reported “recognised academic, vocational, clerical, business or commercial qualifications” asked at each wave  **Responses & coding**:  **0:** No academic qualification **1:** GCDS D-E, GCSE A-C, CSES 2-5, Other Scottish qualifications, Good O levels Scottish standards **2**: As levels or 1 A level; 2+ A levels, Scottish higher/6^th^, diploma **3**: Degree level, Higher degree | N/A | N/A | **Question**: “Please select your highest educational qualification from the list below”  **Responses & coding**:  0: lower education (primary school), lower pre-vocational education (low)  1: pre-vocational education (moderate), secondary education (moderate)  2: middle-level applied education (moderate)  3: higher professional education (high),  university (high) | **Question**: “What is your highest completed educational level?”  **Responses & coding**:  0: None, Uncompleted primary educational level, Primary educational level  1: Lower vocational education, Intermediate general secondary education  2: Intermediate vocational education, Higher general secondary education, Higher vocational education  3: University education |
| *Diet, fruit and vegetable intake* | Fruit intake  0: low  1: low-moderate  2: moderate; high  3: high  Vegetables intake  0: low  1: low-moderate  2: moderate; high  3: high | Fruit: Usual number of servings (portions) eaten per day:  1=I don’t eat fruit  2=Less than 1 per day  3= 1 piece of fruit per day  4=2 pieces of fruit per day  5=3 pieces per day  6= 4 or more per day  **Coding:**  1 = 0  2,3 = 1  4, 5 = 2  6 = 3  Vegetables: Usual number of servings (portions) eaten per day:  1= None  2= Less than one serve  3= 1 serve  4= 2 serves  5= 3 serves  6= 4 serves  7= 5 serves or more  **Coding**:  1, 2 = 0  3,4 = 1  5,6 = 2  7 = 3 | N/A | Fruit: Usual frequency of eating:  1=Every day  2=3-4 times per week  3=1-2 times per week  4=Rarely  Vegetables: Usual frequency of eating:  1=Every day  2=3-4 times per week  3=1-2 times per week  4=Rarely | Vegetables: Frequency of eating in previous 7 days (same for both variables):  0 = not at all  1 = on 1-2 days  2 = on 3-5 days  3 = on 6-7 days | N/A | Fruit Derived from FFQ with 1-year reference period; Continuous variable, g/day.  **Coding**: quartiles  Vegetables: Derived from FFQ with 1-year reference period; Continuous variable, g/day.  **Coding**: quartiles |
| *Self-rated health* | Five-point Likert scale  (note due to country and language-level differences, the five-point categories were not renamed) | **Question**: “In general, how would you say your health is?**”**  **Responses:** excellent, very good, good, fair, poor | **Question**: “ In general, would you say your health is…”  **Responses:** excellent, very good, good, fair, poor | **Question**: “How will you rate your overall health?”  **Reponses**: Very good, good, fairly good, poor, very poor | **Question**: “How would you rate your overall health?”  **Reponses**: Good, rather good, average, rather poor, poor | **Question**: “How could you describe your health status?”  **Question**: Excellent, good, fair, moderate, poor | **Question**: “ In general, would you say your health is…”  **Responses:** excellent, very good, good, fair, poor |
| *Physical function* | Continuous score from 0 to 100 of the SF 10-item physical function, where 0 indicates poor physical function and 100 indicates high physical function. | Available in ALSWH, BCS70, NES and TMS only.  Each used the SF-36 scale. The 10-items included limitations in: vigorous activities, moderate activities, lifting and carrying groceries, climbing several flights of stairs, climbing one flight of stairs, bending, kneeling or stooping, walking about two kilometers, walking about a half kilometer, in walking about 100 metres, in bathing or dressing. Each item had three possible responses: Yes, limited a lot (0); Yes, limited a little (50); No, not limited at all (100). Physical function score was calculated as the average score across all ten items. | | | | | |

**Supplemental Table 5**. Baseline characteristics of participants stratified by sleep regularity index (SRI) group.

| **Baseline characteristic** | **Regular**  **(SRI>87.3)** | **Slightly irregular**  **(SRI 71.6-87.3)** | **Irregular**  **(SRI<71.6)** | **Full sample** |
| --- | --- | --- | --- | --- |
| Sample size | 2,863 | 8,121 | 3,101 | 14,085 |
| **Age** (mean (SD)) | 56.3 (10.1) | 54.2 (9.4) | 52.6 (8.9) | 54.3 (9.5) |
| **Sex** = Female, n (%) | 1,567 (54.7) | 4,569 (56.3) | 1,596 (51.5) | 7,732 (54.9) |
| **Cohort,** n (%) |  |  |  |  |
| ALSWH | 187 (6.5) | 543 (6.7) | 188 (6.1) | 918 (6.5) |
| BCS70 | 889 (31.1) | 2,681 (33.0) | 1,216 (39.2) | 4,786 (34.0) |
| DPhacto | 27 (0.9) | 230 (2.8) | 169 (5.4) | 426 (3.0) |
| FIREA | 33 (1.2) | 92 (1.1) | 22 (0.7) | 147 (1.0) |
| NES | 194 (6.8) | 280 (3.4) | 61 (2.0) | 535 (3.8) |
| TMS | 1,533 (53.5) | 4,295 (52.9) | 1,445 (46.6) | 7,273 (51.6) |
| **Alcohol consumption**^7^**,** n (%) | | | | |
| Tertile 1 | 847 (33.8) | 2,403 (33.8) | 897 (34.8) | 4,147 (34.0) |
| Tertile 2 | 867 (34.6) | 2,435 (34.3) | 843 (32.7) | 4,145 (34.0) |
| Tertile 3 | 790 (31.5) | 2,271 (31.9) | 840 (32.6) | 3,901 (32.0) |
| **Smoking** = Current smoker, n (%) | 268 (9.4) | 1,053 (13.0) | 654 (21.2) | 1,975 (14.1) |
| **Education**, n (%) | | | | |
| None or less than high school | 257 (9.2) | 833 (10.8) | 435 (15.2) | 1,525 (11.4) |
| High school (∼16 years) | 724 (26.0) | 2,062 (26.8) | 897 (31.3) | 3,683 (27.6) |
| Further education (∼16–18 years) | 1,067 (38.4) | 3,122 (40.5) | 1,015 (35.4) | 5,204 (39.0) |
| University degree or higher | 732 (26.3) | 1,683 (21.9) | 519 (18.1) | 2,934 (22.0) |
| **Self-rated health**, n (%) |  |  |  |  |
| Excellent | 427 (15.1) | 971 (12.2) | 268 (8.8) | 1,666 (12.0) |
| Very good | 964 (34.2) | 2,623 (32.9) | 842 (27.6) | 4,429 (32.0) |
| Good | 1,172 (41.5) | 3,449 (43.3) | 1,335 (43.8) | 5,956 (43.0) |
| Fair | 243 (8.6) | 816 (10.2) | 474 (15.6) | 1,533 (11.1) |
| Poor | 16 (0.6) | 112 (1.4) | 128 (4.2) | 256 (1.8) |
| **Medication use**^1^, n (%) | 820 (30.4) | 2,313 (29.9) | 964 (33.1) | 4,097 (30.7) |
| **Prevalent CVD**, n (%) | 276 (9.7) | 789 (9.8) | 339 (11.0) | 1,404 (10.0) |
| **MVPA** mins per day (mean (SD)) ^5^ | 79.7 (29.3) | 75.0 (27.9) | 64.7 (28.4) | 73.7 (28.7) |
| **Diet^6^**, n (%) |  |  |  |  |
| Low | 597 (36.3) | 1,892 (39.8) | 730 (43.7) | 3,219 (39.9) |
| Medium | 677 (41.2) | 1,795 (37.8) | 601 (36.0) | 3,073 (38.1) |
| High | 370 (22.5) | 1,067 (22.4) | 338 (20.3) | 1,775 (22.0) |
| **Body Mass Index**, kg/m^2^ (mean (SD)) | 26.0 (4.1) | 26.7 (4.4) | 28.0 (4.8) | 26.8 (4.5) |
| **Waist circumference**, cm (mean (SD)) | 91.7 (12.6) | 93.3 (12.9) | 97.0 (13.7) | 93.8 (13.2) |
| **HDL**^2^, mmol/L (mean (SD)) | 1.6 (0.4) | 1.6 (0.4) | 1.5 (0.4) | 1.6 (0.4) |
| **LDL**^3^, mmol/L (mean (SD)) | 3.3 (0.9) | 3.4 (0.9) | 3.4 (1.0) | 3.4 (0.9) |
| **Triglycerides**, mmol/L (mean (SD)) | 1.3 (0.7) | 1.4 (0.8) | 1.6 (0.8) | 1.4 (0.8) |
| **HbA1C**^4^, mmol/mol (mean (SD)) | 37.3 (6.2) | 37.5 (6.8) | 38.6 (8.0) | 37.7 (7.0) |
| **Systolic blood pressure**, mmHg (mean (SD)) | 130.8 (17.5) | 129.4 (16.4) | 129.8 (16.3) | 129.8 (16.6) |
| **Diastolic blood pressure**, mmHg (mean (SD)) | 75.7 (9.9) | 76.4 (9.7) | 77.6 (10.1) | 76.5 (9.8) |
| **Sleep regularity index** (mean (SD)) | 90.2 (1.9) | 80.3 (4.3) | 62.3 (7.6) | 78.4 (10.6) |
| **Sleep duration**, h/day (mean (SD)) | 7.0 (0.9) | 7.1 (1.0) | 7.4 (1.3) | 7.2 (1.0) |
| **Sleep efficiency**, % (mean (SD)) | 90.6 (6.5) | 87.8 (7.6) | 80.7 (10.0) | 86.8 (8.7) |

SRI: Sleep Regularity Index; SD: standard deviation.

^1^Lipid-modifying, hypertensive, and glucose-lowering medications; ^2^High-density lipoprotein cholesterol; ^3^Low-density lipoprotein cholesterol, ^4^Glycated Haemoglobin, ^5^Accelerometer-derived moderate-to-vigorous physical activity, ^6^ Self-reported daily intake of fruits and vegetables served as a proxy for dietary quality by asking, ^7^Self-reported alcohol consumption.

**Supplemental Table 6**. Baseline characteristics of participants stratified by sleep efficiency.

| **Baseline characteristic** | **Low**  **(< 85.3)** | **Medium**  **(85.3-91.8)** | **High**  **(>91.8)** | **Full sample** |
| --- | --- | --- | --- | --- |
| Sample size | 4,695 | 4,695 | 4,695 | 14,085 |
| **Age** (mean (SD)) | 55.0 (9.7) | 54.9 (9.7) | 53.0 (8.9) | 54.3 (9.5) |
| **Sex** = Female, n (%) | 2,382 (50.7) | 2,635 (56.1) | 2,715 (57.8) | 7,732 (54.9) |
| **Cohort,** n (%) |  |  |  |  |
| ALSWH | 268 (5.7) | 313 (6.7) | 337 (7.2) | 918 (6.5) |
| BCS70 | 1,480 (31.5) | 1,519 (32.4) | 1,787 (38.1) | 4,786 (34.0) |
| DPhacto | 152 (3.2) | 150 (3.2) | 124 (2.6) | 426 (3.0) |
| FIREA | 44 (0.9) | 48 (1.0) | 55 (1.2) | 147 (1.0) |
| NES | 179 (3.8) | 202 (4.3) | 154 (3.3) | 535 (3.8) |
| TMS | 2,572 (54.8) | 2,463 (52.5) | 2,238 (47.7) | 7,273 (51.6) |
| **Alcohol consumption**^7^**,** n (%) | | | | |
| Tertile 1 | 1,391 (34.9) | 1,357 (33.1) | 1,399 (34.1) | 4,147 (34.0) |
| Tertile 2 | 1,285 (32.2) | 1,420 (34.6) | 1,440 (35.1) | 4,145 (34.0) |
| Tertile 3 | 1,315 (32.9) | 1,325 (32.3) | 1,261 (30.8) | 3,901 (32.0) |
| **Smoking** = Current smoker, n (%) | 785 (16.8) | 610 (13.1) | 580 (12.4) | 1,975 (14.1) |
| **Education**, n (%) | | | | |
| None or less than high school | 584 (13.2) | 476 (10.7) | 465 (10.4) | 1,525 (11.4) |
| High school (∼16 years) | 1,349 (30.5) | 1,214 (27.3) | 1,120 (25.1) | 3,683 (27.6) |
| Further education (∼16–18 years) | 1,681 (38.0) | 1,797 (40.4) | 1,726 (38.6) | 5,204 (39.0) |
| University degree or higher | 814 (18.4) | 963 (21.6) | 1,157 (25.9) | 2,934 (22.0) |
| **Self-rated health**, n (%) |  |  |  |  |
| Excellent | 407 (8.8) | 573 (12.4) | 686 (14.8) | 1,666 (12.0) |
| Very good | 1,248 (27.1) | 1,558 (33.7) | 1,623 (35.1) | 4,429 (32.0) |
| Good | 2,121 (46.1) | 1,957 (42.4) | 1,878 (40.6) | 5,956 (43.0) |
| Fair | 682 (14.8) | 466 (10.1) | 385 (8.3) | 1,533 (11.1) |
| Poor | 141 (3.1) | 64 (1.4) | 51 (1.1) | 256 (1.8) |
| **Medication use**^1^, n (%) | 1,668 (37.6) | 1,349 (30.4) | 1,080 (24.1) | 4,097 (30.7) |
| **Prevalent CVD**, n (%) | 535 (11.5) | 493 (10.6) | 376 (8.1) | 1,404 (10.0) |
| **MVPA** mins per day (mean (SD)) ^5^ | 69.4 (29.5) | 75.0 (28.3) | 76.7 (27.9) | 73.7 (28.7) |
| **Diet^6^**, n (%) |  |  |  |  |
| Low | 1,139 (40.9) | 1,056 (38.6) | 1,024 (40.3) | 3,219 (39.9) |
| Medium | 1,043 (37.4) | 1,048 (38.3) | 982 (38.6) | 3,073 (38.1) |
| High | 605 (21.7) | 632 (23.1) | 538 (21.1) | 1,775 (22.0) |
| **Body Mass Index**, kg/m^2^ (mean (SD)) | 27.8 (4.8) | 26.7 (4.3) | 26.1 (4.1) | 26.8 (4.5) |
| **Waist circumference**, cm (mean (SD)) | 96.9 (13.6) | 93.2 (12.9) | 91.3 (12.3) | 93.8 (13.2) |
| **HDL**^2^, mmol/L (mean (SD)) | 1.5 (0.4) | 1.6 (0.4) | 1.6 (0.4) | 1.6 (0.4) |
| **LDL**^3^, mmol/L (mean (SD)) | 3.4 (0.9) | 3.4 (0.9) | 3.4 (0.9) | 3.4 (0.9) |
| **Triglycerides**, mmol/L (mean (SD)) | 1.5 (0.8) | 1.4 (0.8) | 1.4 (0.7) | 1.4 (0.8) |
| **HbA1C**^4^, mmol/mol (mean (SD)) | 38.9 (7.9) | 37.6 (6.8) | 36.7 (6.0) | 37.7 (7.0) |
| **Systolic blood pressure**, mmHg (mean (SD)) | 131.6 (16.5) | 130.0 (16.8) | 127.8 (16.4) | 129.8 (16.6) |
| **Diastolic blood pressure**, mmHg (mean (SD)) | 77.6 (9.9) | 76.4 (9.8) | 75.5 (9.7) | 76.5 (9.8) |
| **Sleep regularity index** (mean (SD)) | 72.8 (12.1) | 79.4 (8.9) | 82.9 (7.5) | 78.4 (10.6) |
| **Sleep duration**, h/day (mean (SD)) | 6.9 (1.2) | 7.3 (0.9) | 7.4 (0.8) | 7.2 (1.0) |
| **Sleep efficiency**, % (mean (SD)) | 76.9 (7.3) | 88.8 (1.9) | 94.8 (2.0) | 86.8 (8.7) |

SRI: Sleep Regularity Index; SD: standard deviation.

^1^Lipid-modifying, hypertensive, and glucose-lowering medications; ^2^High-density lipoprotein cholesterol; ^3^Low-density lipoprotein cholesterol, ^4^Glycated Haemoglobin, ^5^Accelerometer-derived moderate-to-vigorous physical activity, ^6^ Self-reported daily intake of fruits and vegetables served as a proxy for dietary quality by asking, ^7^Self-reported alcohol consumption.

**Supplemental Table 7**. Multiplicative interaction between sleep parameters and sex.

| Factors | Coefficient (β) | 95% CI | P-value |
| --- | --- | --- | --- |
| **Sleep duration x Sex** | | | |
| Short x Male | ref |  |  |
| Adequate x Female | -0.03 | -0.09, 0.04 | 0.424 |
| Long x Female | -0.04 | -0.10, 0.06 | 0.579 |
| **Sleep regularity x Sex** | | | |
| Regular x Male | ref |  |  |
| Slightly irregular x Female | 0.05 | -0.03, 0.12 | 0.210 |
| Irregular x Female | 0.02 | -0.07, 0.11 | 0.682 |
| **Sleep efficiency x Sex** | | | |
| Low x Male | ref |  |  |
| Medium x Female | -0.04 | -0.11, 0.03 | 0.305 |
| High x Female | -0.12 | -0.19, -0.05 | <0.001 |

**Supplemental Table 8. Interaction between sleep parameters and age.**

| Factors | Coefficient (β) | 95% CI | P-value |
| --- | --- | --- | --- |
| **Sleep duration x age** | | | |
| Short x age | ref |  |  |
| Adequate x age | 0.01 | 0.001, 0.007 | 0.020 |
| Long x age | 0.01 | 0.001, 0.008 | 0.069 |
| **Sleep regularity x age** | | | |
| Regular x age | ref |  |  |
| Slightly irregular x age | -0.002 | -0.005, 0.005 | 0.225 |
| Irregular x age | -0.001 | -0.004, 0.005 | 0.933 |
| **Sleep efficiency x age** | | | |
| Regular x age | ref |  |  |
| Slightly irregular x age | 0.002 | -0.002, 0.005 | 0.432 |
| Irregular x age | 0.007 | 0.003, 0.011 | <0.001 |

**Supplemental Figure 1**. Association of sleep duration with cardiometabolic health markers including A) BMI, B) waist circumference, C) HDL cholesterol, D) LDL cholesterol, E) Triglycerides, F) Glycated haemoglobin (HbA1c), G) Systolic and H) Diastolic blood pressure.

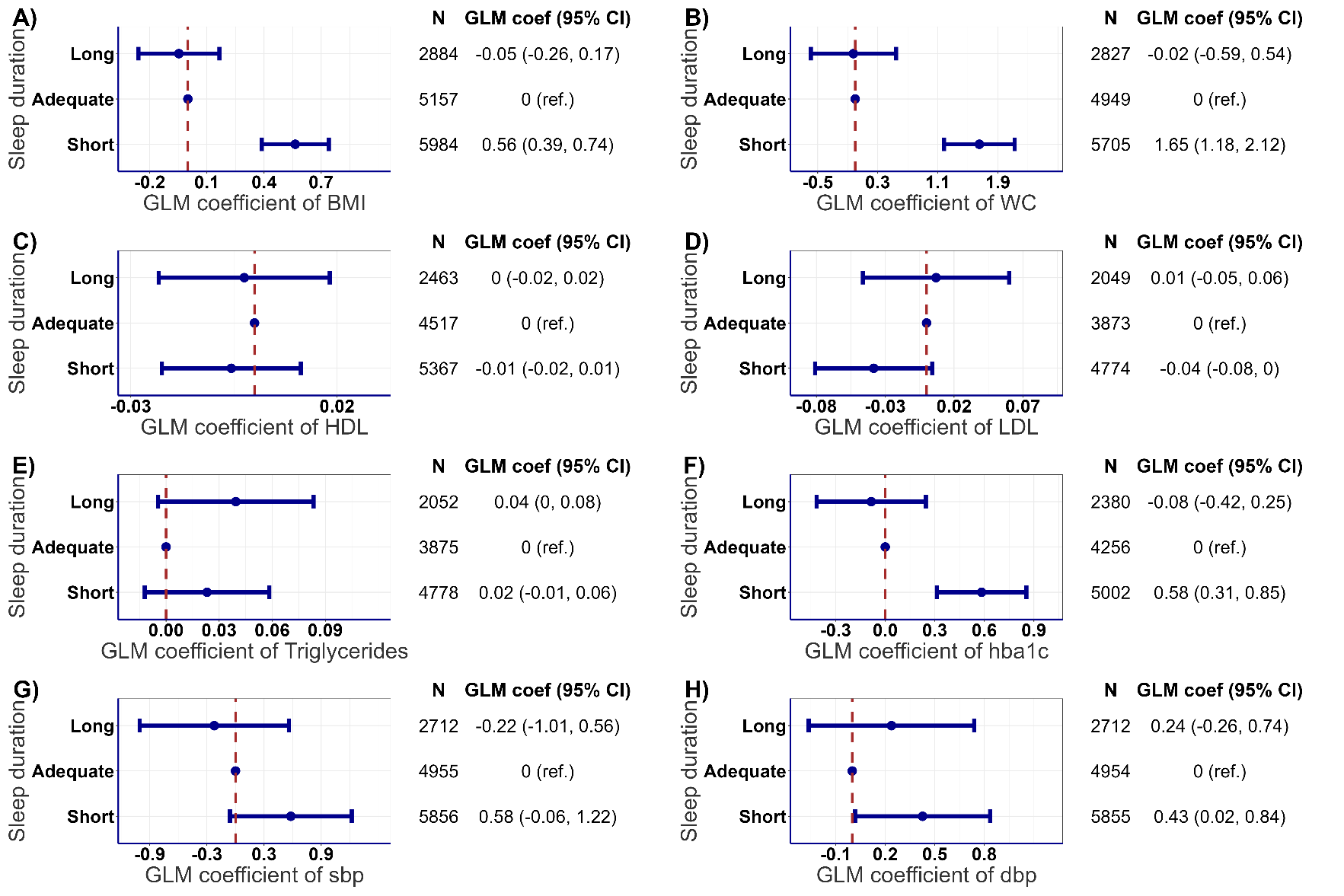

**Legend:** Sleep duration was categorized into short (<7h/day), adequate (7-8h/day) and long (>8h/day). Reference group was set to ‘adequate’ for sleep duration. All models were adjusted for age, sex, cohort, smoking, alcohol consumption, medication use, previous cardiovascular incidence, moderate-to-vigorous physical activity, sleep regularity and sleep efficiency. N=14,025 (BMI, kg/m2); 13,481 (waist circumference, cm); 12,347 (HDL, mmol/L); 10,696 (LDL, mmol/L); 10,705 (triglycerides, mmol/L); 11,638 (HbA1c, mmol/mol); 13,523 (SBP, mmol/mol); 13,521 (DBP, mmol/mol). GLM coefficients represent the mean differences between the reference group and each of the other groups. SRI: sleep regularity index, BMI: body mass index, SBP: systolic blood pressure, DBP: diastolic blood pressure.

**Supplemental Figure 2**. Association of sleep regularity with cardiometabolic health markers including A) BMI, B) waist circumference, C) HDL cholesterol, D) LDL cholesterol, E) Triglycerides, F) Glycated haemoglobin (HbA1c), G) Systolic and H) Diastolic blood pressure.

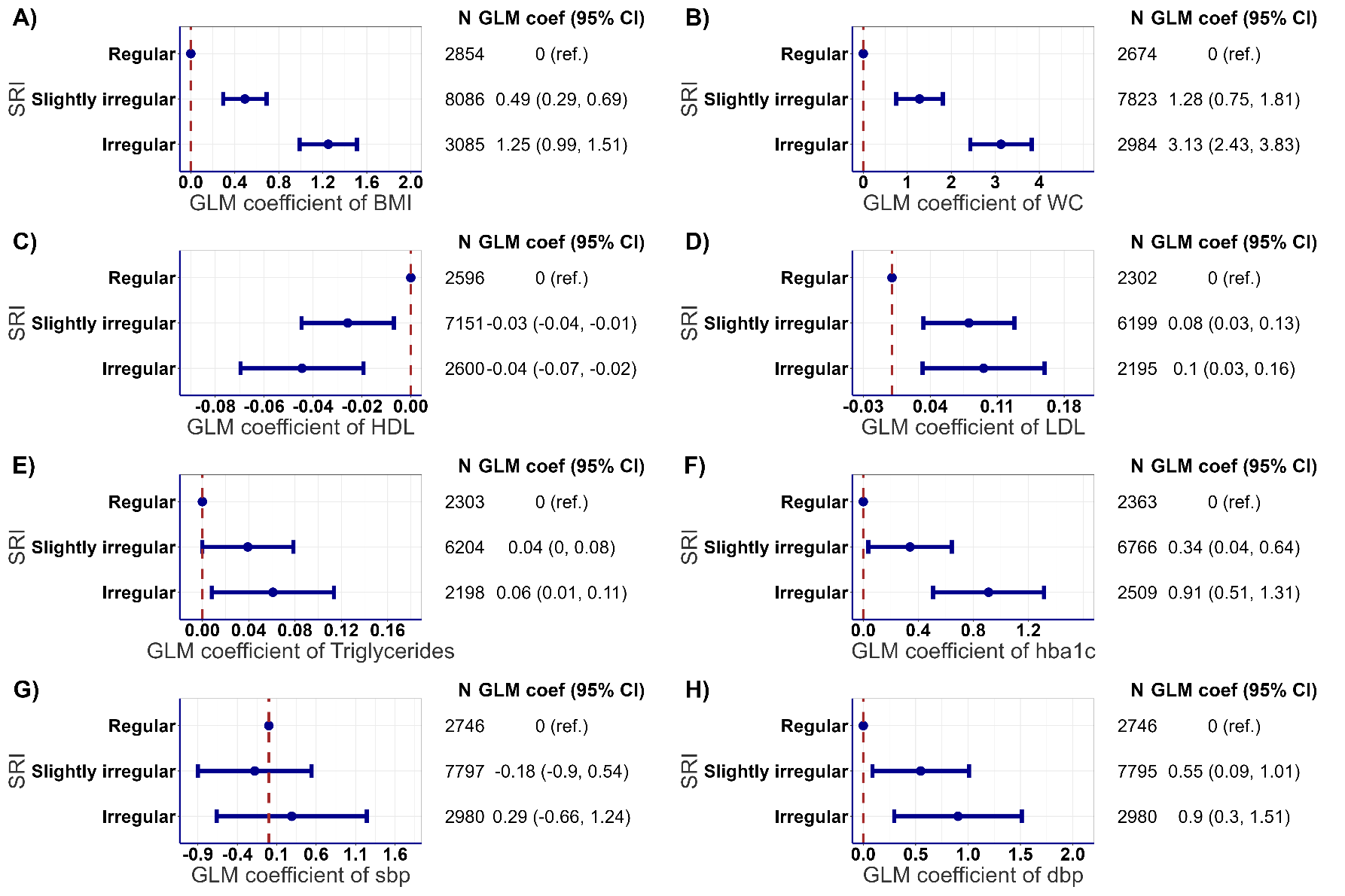

**Legend:** Sleep regularity was categorized into regular (SRI> 87.3), slightly irregular (71.6≤SRI≤ 87.3), and irregular (SRI<71.6). Reference group was set to regular for sleep regularity. All models were adjusted for age, sex, cohort, smoking, alcohol consumption, medication use, previous cardiovascular incidence, moderate-to-vigorous physical activity, sleep duration and sleep efficiency. N=14,025 (BMI, kg/m2); 13,481 (waist circumference, cm); 12,347 (HDL, mmol/L); 10,696 (LDL, mmol/L); 10,705 (triglycerides, mmol/L); 11,638 (HbA1c, mmol/mol); 13,523 (SBP, mmol/mol); 13,521 (DBP, mmol/mol). GLM coefficients represent the mean differences between the reference group and each of the other groups. SRI: sleep regularity index, BMI: body mass index, SBP: systolic blood pressure, DBP: diastolic blood pressure.

**Supplemental Figure 3**. Association of sleep efficiency with cardiometabolic health markers including A) BMI, B) waist circumference, C) HDL cholesterol, D) LDL cholesterol, E) Triglycerides, F) Glycated haemoglobin (HbA1c), G) Systolic and H) Diastolic blood pressure.

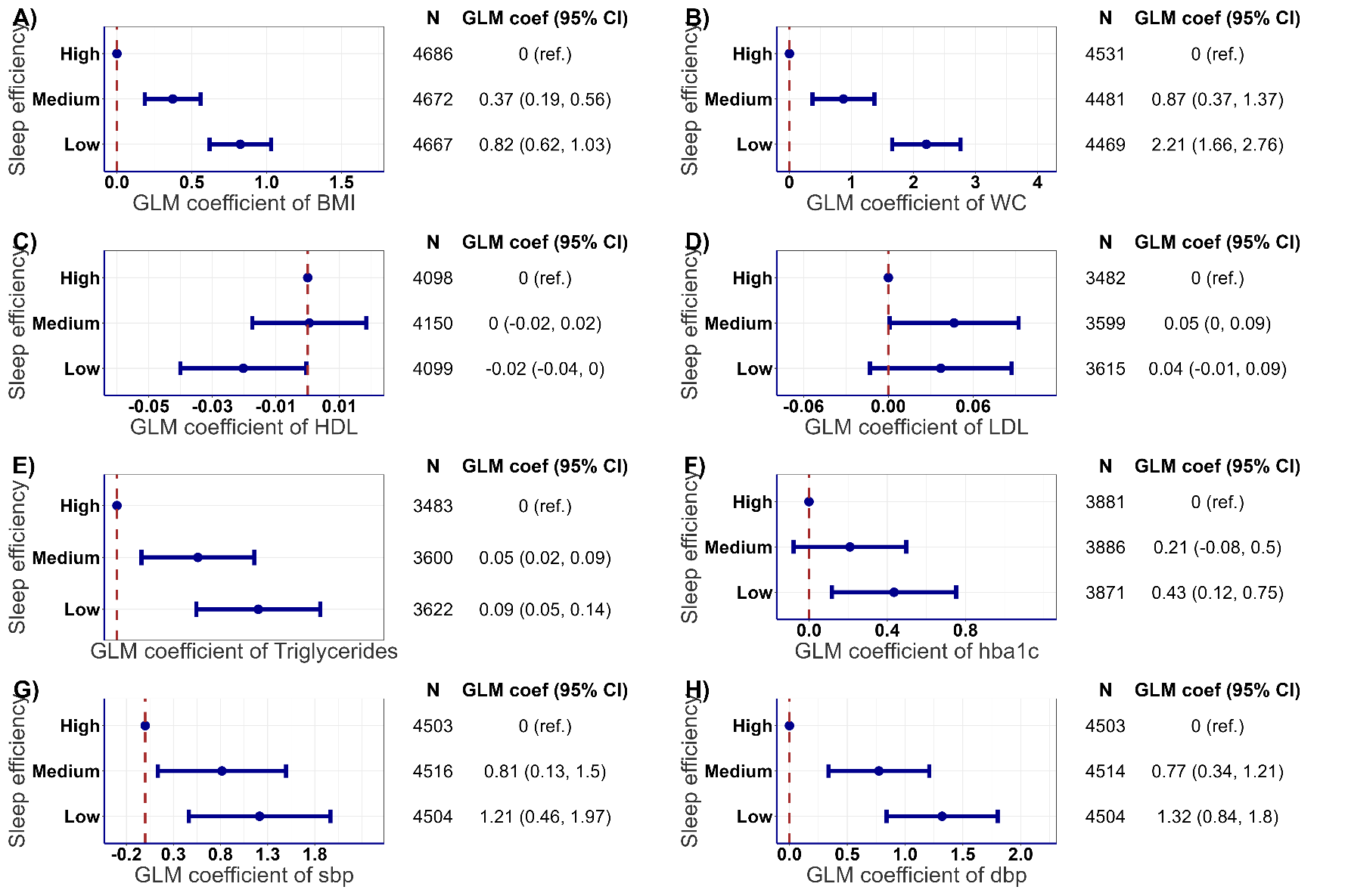

**Legend:** Sleep efficiency was categorized into high efficiency (>91.8), medium (85.3≤ efficiency ≤ 91.8), and low (<85.3). Reference group was set to high for sleep efficiency. All models were adjusted for age, sex, cohort, smoking, alcohol consumption, medication use, previous cardiovascular incidence, moderate-to-vigorous physical activity, sleep duration and sleep regularity. N=14,025 (BMI, kg/m2); 13,481 (waist circumference, cm); 12,347 (HDL, mmol/L); 10,696 (LDL, mmol/L); 10,705 (triglycerides, mmol/L); 11,638 (HbA1c, mmol/mol); 13,523 (SBP, mmol/mol); 13,521 (DBP, mmol/mol). GLM coefficients represent the mean differences between the reference group and each of the other groups. SRI: sleep regularity index, BMI: body mass index, SBP: systolic blood pressure, DBP: diastolic blood pressure.

**Supplemental Figure 4**. Joint association of sleep duration, regularity and efficiency with composite cardiometabolic risk score with additional adjustment for diet and education.

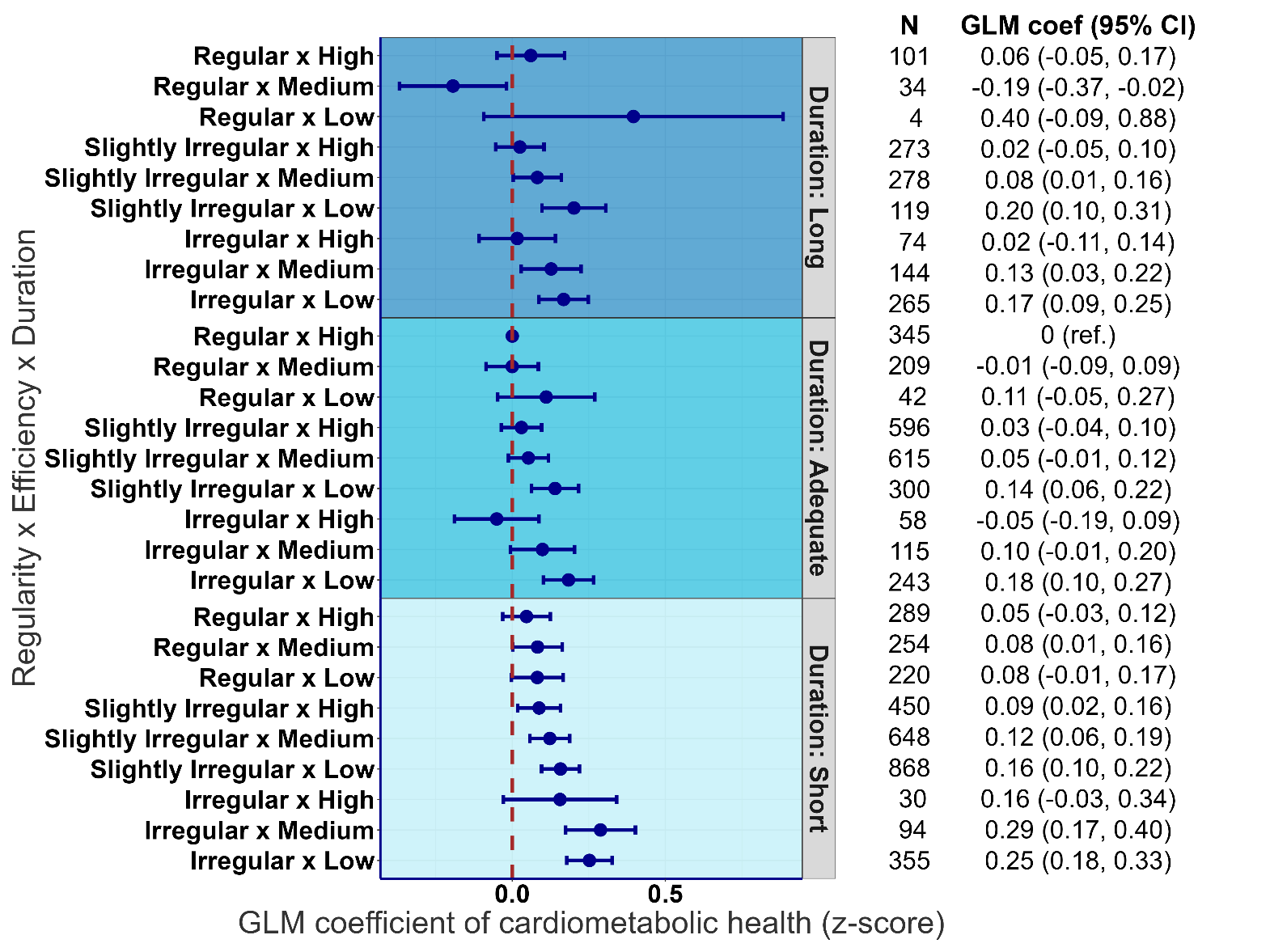

**Legend:** Sleep duration was categorized into short (<7h/day), adequate (7-8h/day) and long (>8h/day), sleep regularity (irregular, SRI<71.6; slightly irregular, 71.6≤SRI≤ 87.3; regular, SRI> 87.3) and sleep efficiency (low, efficiency<85.3; medium, 85.3≤ efficiency ≤ 91.8; high, efficiency >91.8) were categorized into tertiles. Reference group was set to short duration, irregular and low efficiency of sleep. All models were adjusted for age, sex, smoking, alcohol consumption, medication use, previous cardiovascular incidence, diet, education and moderate-to-vigorous physical activity. N= 7,023. GLM coefficients represent the mean differences between the reference group and each of the other groups. Cohort was not adjusted as not all cohorts had information on diet and education.

**Supplemental Figure 5**. Joint association of sleep duration, regularity and efficiency with composite cardiometabolic risk score with additional adjustment for body fat percentage.

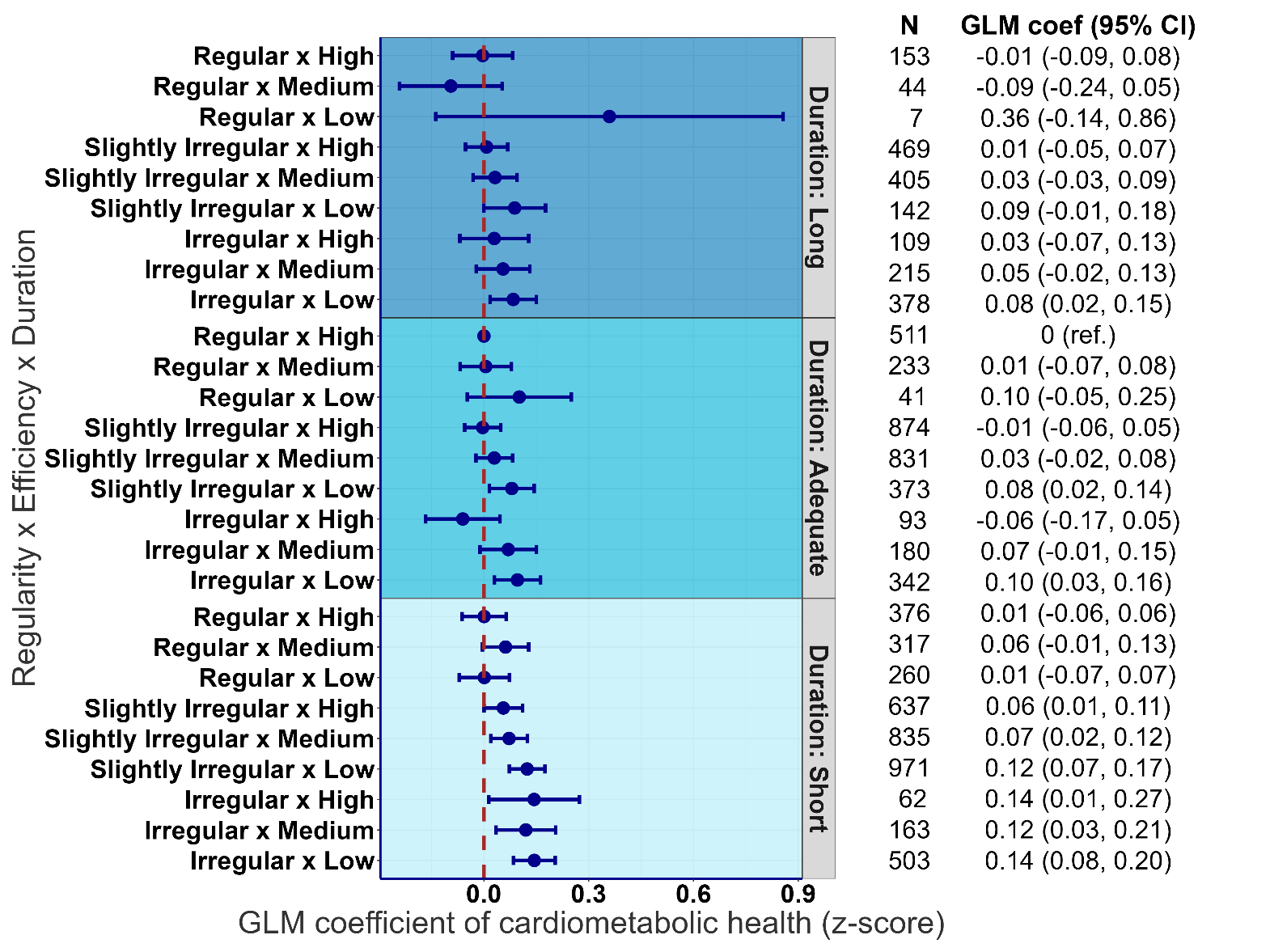

**Legend:** Sleep duration was categorized into short (<7h/day), adequate (7-8h/day) and long (>8h/day), sleep regularity (irregular, SRI<71.6; slightly irregular, 71.6≤SRI≤ 87.3; regular, SRI> 87.3) and sleep efficiency (low, efficiency<85.3; medium, 85.3≤ efficiency ≤ 91.8; high, efficiency >91.8) were categorized into tertiles. Reference group was set to short duration, irregular and low efficiency of sleep. All models were adjusted for age, sex, cohort, smoking, alcohol consumption, medication use, previous cardiovascular incidence, body fat percentage, and moderate-to-vigorous physical activity. N= 9,524. GLM coefficients represent the mean differences between the reference group and each of the other groups. Only 1970 British Cohort Study and The Maastricht Study had data on body fat percentage.

**Supplemental Figure 6**. Joint association of sleep duration, regularity and efficiency with composite cardiometabolic risk score with additional adjustment for physical function.

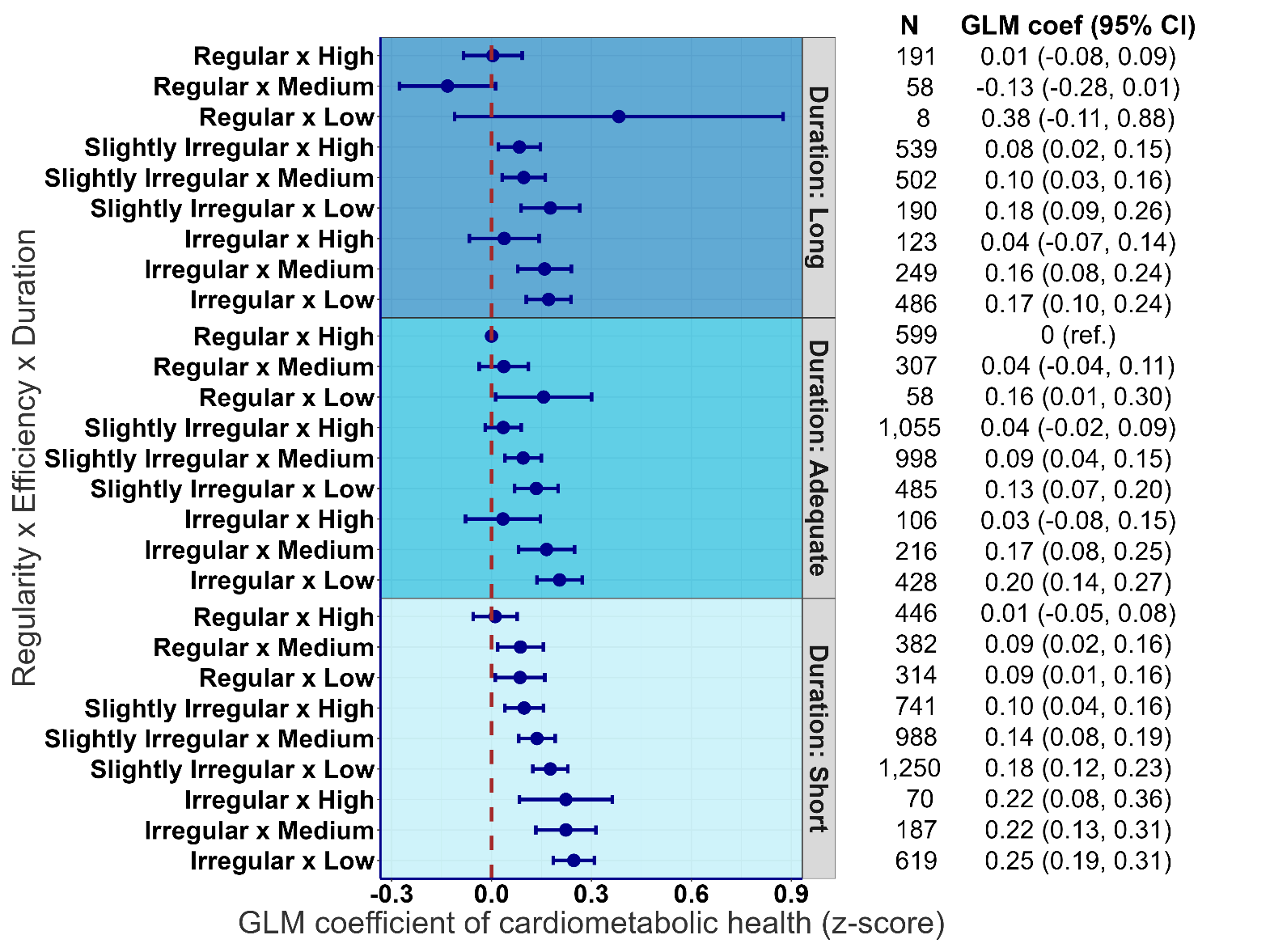

**Legend:** Sleep duration was categorized into short (<7h/day), adequate (7-8h/day) and long (>8h/day), sleep regularity (irregular, SRI<71.6; slightly irregular, 71.6≤SRI≤ 87.3; regular, SRI> 87.3) and sleep efficiency (low, efficiency<85.3; medium, 85.3≤ efficiency ≤ 91.8; high, efficiency >91.8) were categorized into tertiles. Reference group was set to short duration, irregular and low efficiency of sleep. All models were adjusted for age, sex, cohort, smoking, alcohol consumption, medication use, previous cardiovascular incidence, physical function and moderate-to-vigorous physical activity. N= 11,595. GLM coefficients represent the mean differences between the reference group and each of the other groups.

**Supplemental Figure 7**. Joint association of sleep duration, regularity and efficiency with composite cardiometabolic risk score excluding those who self-reported health as poor or fair.

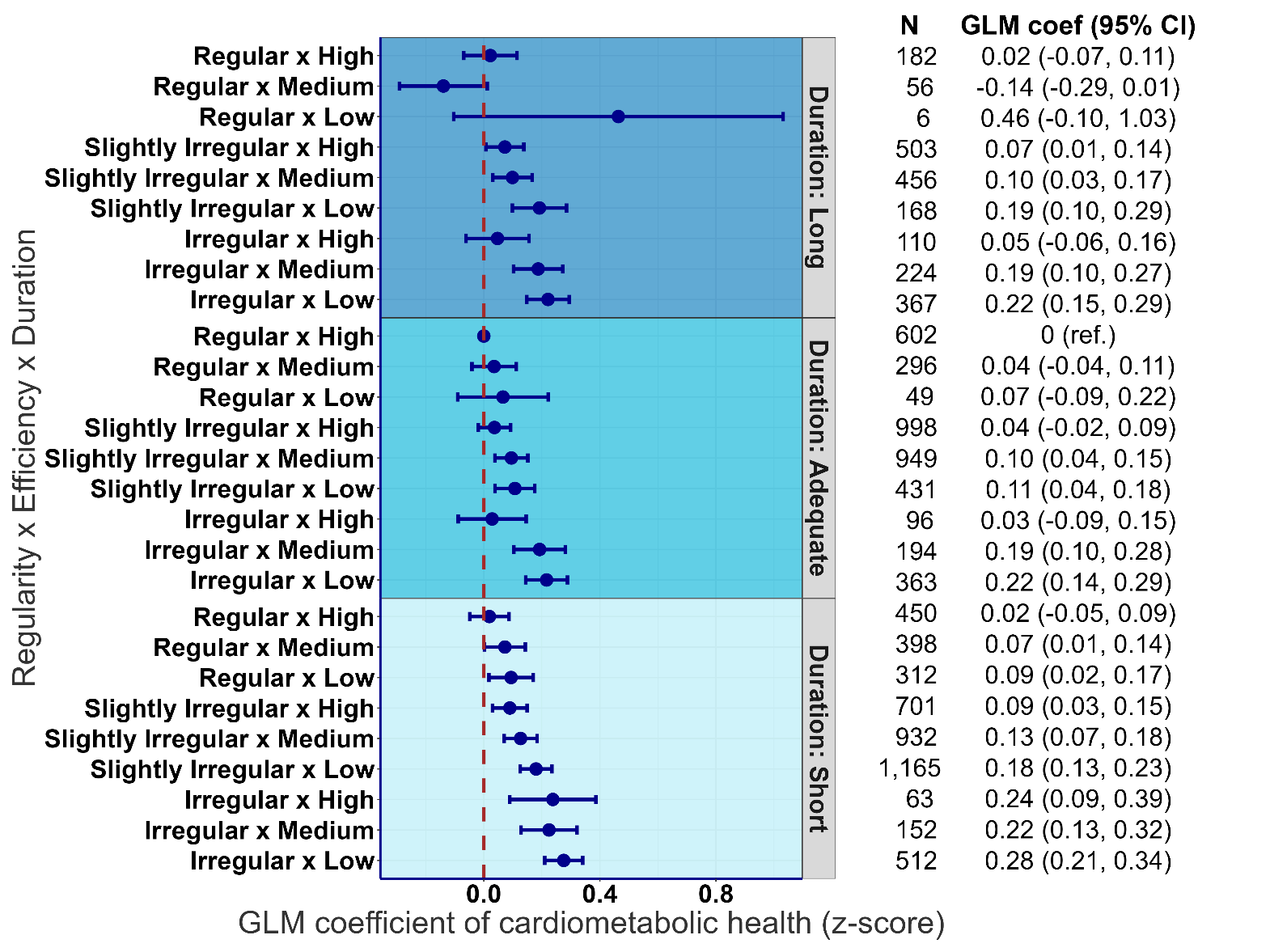

**Legend:** Sleep duration was categorized into short (<7h/day), adequate (7-8h/day) and long (>8h/day), sleep regularity (irregular, SRI<71.6; slightly irregular, 71.6≤SRI≤ 87.3; regular, SRI> 87.3) and sleep efficiency (low, efficiency<85.3; medium, 85.3≤ efficiency ≤ 91.8; high, efficiency >91.8) were categorized into tertiles. Reference group was set to short duration, irregular and low efficiency of sleep. All models were adjusted for age, sex, cohort, smoking, alcohol consumption, medication use, previous cardiovascular incidence and moderate-to-vigorous physical activity. N= 10,735. GLM coefficients represent the mean differences between the reference group and each of the other groups.

**Supplemental Figure 8**. Joint association of sleep duration, regularity and efficiency with composite cardiometabolic risk score excluding those with previous history of cardiovascular disease.

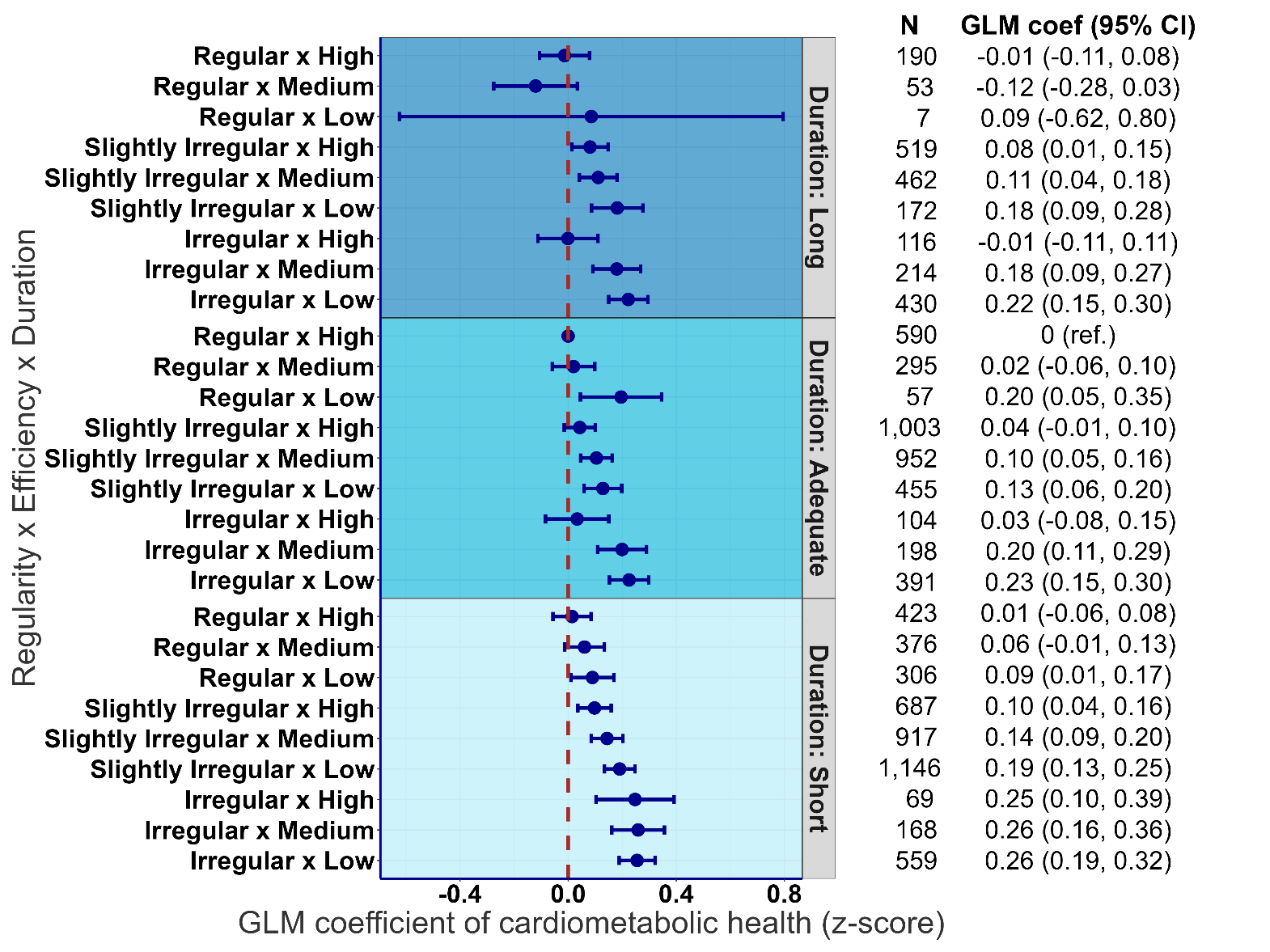

**Legend:** Sleep duration was categorized into short (<7h/day), adequate (7-8h/day) and long (>8h/day), sleep regularity (irregular, SRI<71.6; slightly irregular, 71.6≤SRI≤ 87.3; regular, SRI> 87.3) and sleep efficiency (low, efficiency<85.3; medium, 85.3≤ efficiency ≤ 91.8; high, efficiency >91.8) were categorized into tertiles. Reference group was set to short duration, irregular and low efficiency of sleep. All models were adjusted for age, sex, cohort, smoking, alcohol consumption, medication use, and moderate-to-vigorous physical activity. N= 10,859. GLM coefficients represent the mean differences between the reference group and each of the other groups.

**Supplemental Figure 9**. Joint association of sleep duration, regularity and efficiency with composite cardiometabolic risk score excluding those with BMI over 35.

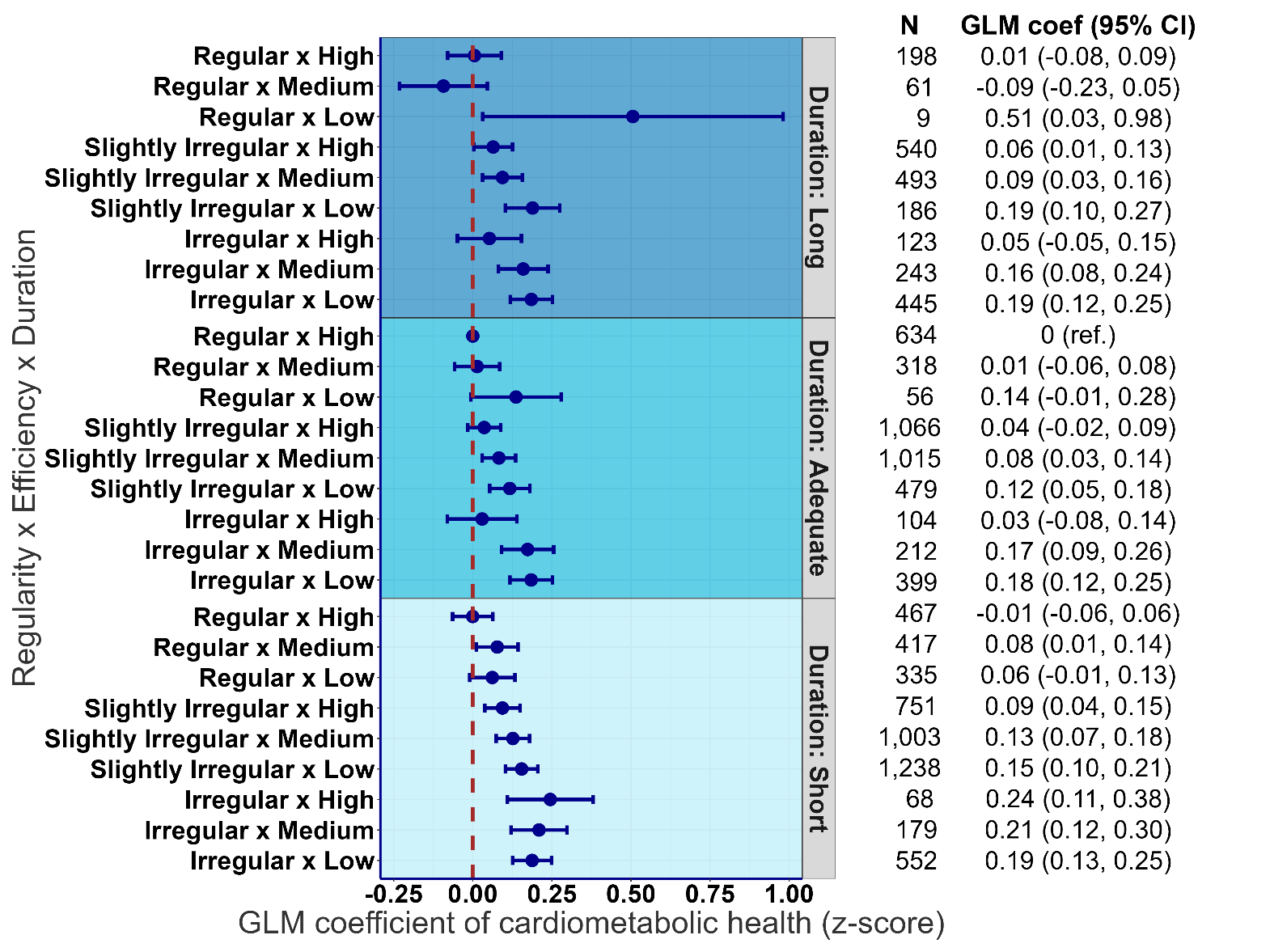

**Legend:** Sleep duration was categorized into short (<7h/day), adequate (7-8h/day) and long (>8h/day), sleep regularity (irregular, SRI<71.6; slightly irregular, 71.6≤SRI≤ 87.3; regular, SRI> 87.3) and sleep efficiency (low, efficiency<85.3; medium, 85.3≤ efficiency ≤ 91.8; high, efficiency >91.8) were categorized into tertiles. Reference group was set to short duration, irregular and low efficiency of sleep. All models were adjusted for age, sex, cohort, smoking, alcohol consumption, medication use, and moderate-to-vigorous physical activity. N= 11,591. GLM coefficients represent the mean differences between the reference group and each of the other groups.

**Supplemental Figure 10.** Three-way Joint association of sleep duration, regularity and efficiency with an alternative composite cardiometabolic risk score.

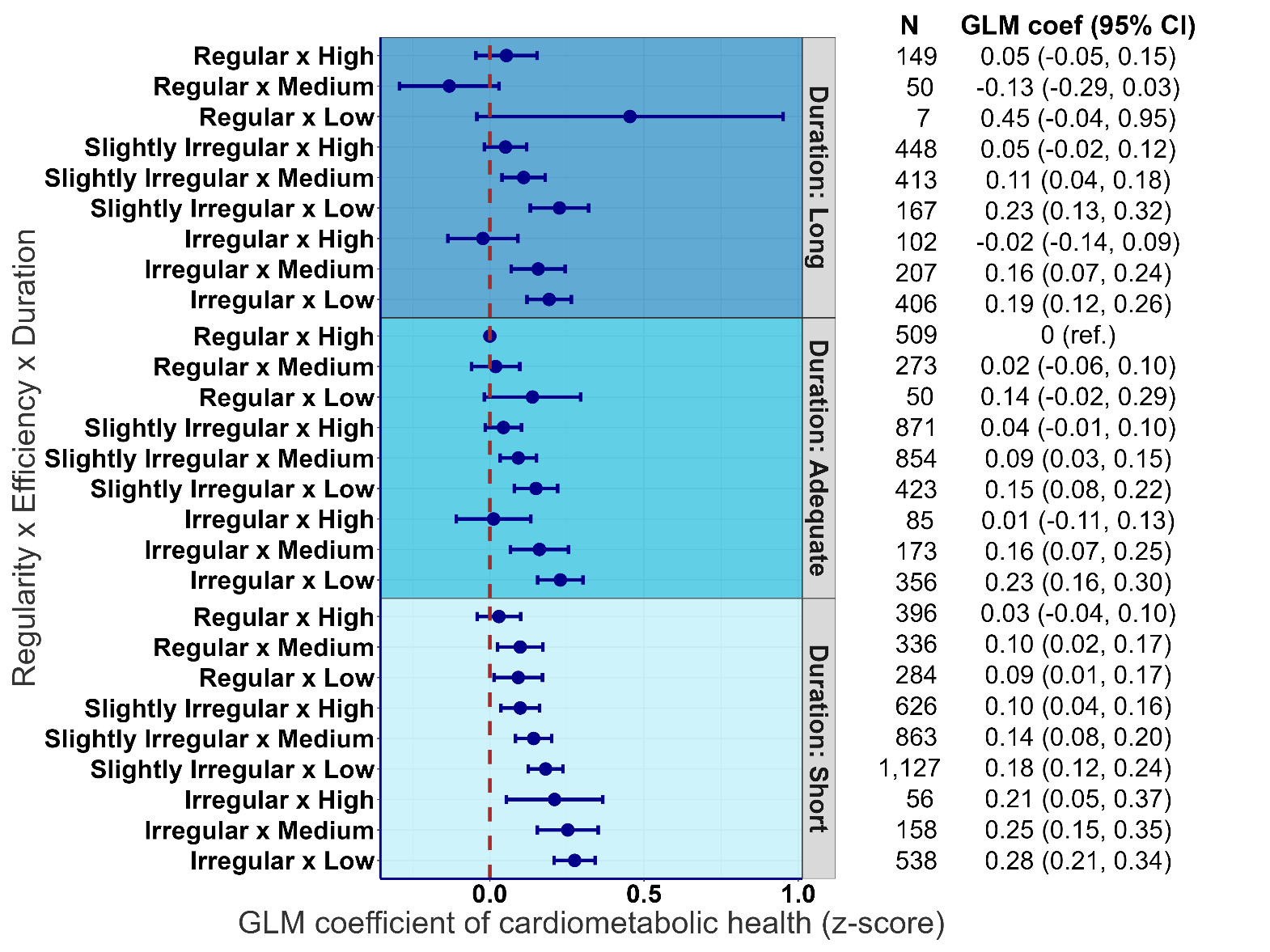

**Legend:** The alternative composite cardiometabolic risk score uses the same set of eight biomarkers but applies a stricter criterion by excluding individuals with missing data in any biomarker, ensuring the score is based solely on complete cases. Sleep duration was categorized into short (<7h/day), adequate (7-8h/day) and long (>8h/day), sleep regularity (irregular, SRI<71.6; slightly irregular, 71.6≤SRI≤ 87.3; regular, SRI>87.3) and sleep efficiency (low, efficiency<85.3; medium, 85.3≤ efficiency ≤ 91.8; high, efficiency >91.8) were categorized into tertiles. Reference group was set to adequate duration with regular and high efficiency of sleep. All models were adjusted for age, sex, cohort, smoking, alcohol consumption, medication use, previous cardiovascular incidence and moderate-to-vigorous physical activity. N= 9,927. GLM coefficients represent the mean differences between the reference group and each of the other groups. SRI: sleep regularity index.

**Supplemental Figure 11.** Three-way Joint association of sleep duration, regularity and efficiency with composite cardiometabolic risk score in participants with at least 7 days of valid wear days (≥20 hours per day).

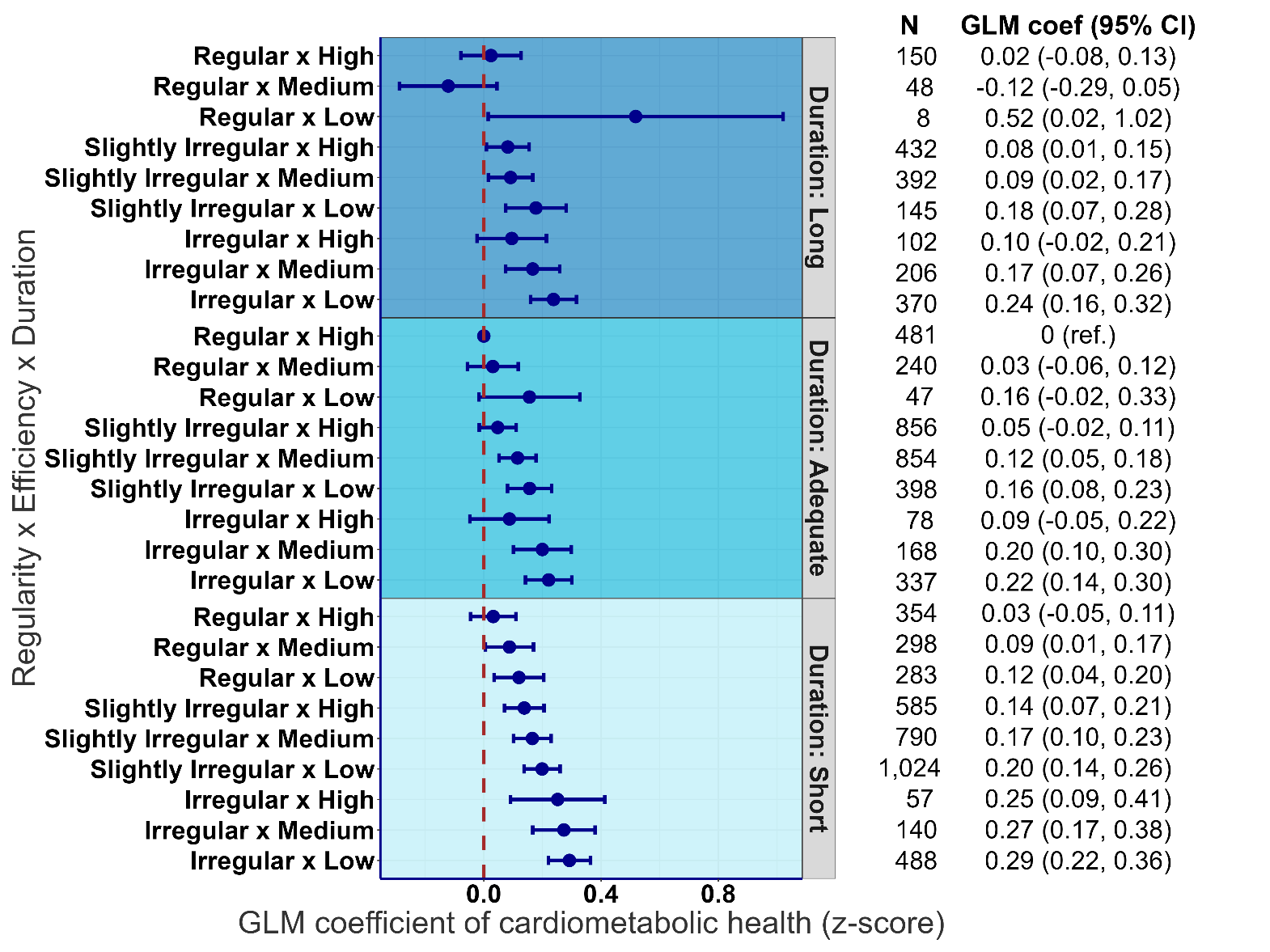

**Legend:** Sleep duration was categorized into short (<7h/day), adequate (7-8h/day) and long (>8h/day), sleep regularity (irregular, SRI<71.6; slightly irregular, 71.6≤SRI≤ 87.3; regular, SRI>87.3) and sleep efficiency (low, efficiency<85.3; medium, 85.3≤ efficiency ≤ 91.8; high, efficiency >91.8) were categorized into tertiles. Reference group was set to adequate duration with regular and high efficiency of sleep. All models were adjusted for age, sex, cohort, smoking, alcohol consumption, medication use, previous cardiovascular incidence and moderate-to-vigorous physical activity. N= 9,331. GLM coefficients represent the mean differences between the reference group and each of the other groups. SRI: sleep regularity index.
